# Accelerated grey matter degeneration in relapsing remitting multiple sclerosis

**DOI:** 10.1101/2025.07.01.25330635

**Authors:** Max Korbmacher, Ingrid Anne Lie, Kristin Wesnes, Eric Westman, Thomas Espeseth, Ole Andreas Andreassen, Hanne Harbo, Gro Owren Nygaard, Lars Tjelta Westlye, Stig Wergeland, Kjell-Morten Myhr, Einar August Høgestøl, Øivind Torkildsen

**Author notes:** These authors contributed equally to this work (Einar August Høgestøl, Øivind Torkildsen). Correspondence to: Max Korbmacher Jonas Lies vei 71, 5053 Bergen, Norway.

## Abstract

**Background:** Grey matter (GM) atrophy has been suggested as the most accurate marker of neurodegeneration in multiple sclerosis (MS) progression. However, long-term comparisons between MS and healthy controls (HC) are rare, and the most significantly atrophying brain regions and their clinical importance still need robust and longitudinal validation.

**Methods:** This multi-cohort longitudinal observational study used two relapsing remitting MS (RRMS) cohorts (N=386, T_1_w-scans=940) sampled for up to 12 years to map grey matter atrophy localisation and disease progression (Expanded Disability Status Scale (EDSS), Paced Auditory Serial Addition Test (PASAT), Fatigue Severity Scale (FSS)). The identified region-specific significant atrophy was compared with 2,163 HCs (T_1_w-scans=4,326).

**Results:** The strongest, replicable, significant brain atrophy patterns in RRMS were found in the frontal lobes, specifically, in the superior frontal cortex (SFC, β_age_≤-0.27), pars orbitalis (β_age_≤-0.25), and thalami (β_age_≤-0.20). Across samples, the examined significant atrophy patterns in MS were more pronounced than in a sample of more than 20-years older HCs in the right SFC (Z>2.41, p<0.009), caudal-middle frontal cortex (Z>2.08, p<0.019), caudate (Z>2.09, p<0.019), and the left frontal pole (Z>2.42, p<0.008). The replicability of associations between volumetric and clinical outcome changes was limited and better described by whole-brain than regional volume changes.

**Conclusions:** Our findings indicate accelerated GM atrophy in people with RRMS, specific to the SFL and thalamus and related to disability-progression. These results can inform further studies examining the role of thalamic and SFC atrophy as MS-biomarkers.

**Key Messages:** *What is already known on this topic:* Grey matter atrophy is considered a hallmark of neurodegeneration in multiple sclerosis, with regional atrophy, especially in the thalamus, linked to disability progression. However, few studies have longitudinally examined region-specific atrophy and its clinical relevance or compared MS-related atrophy with normal ageing using large control samples.

*What this study adds:* This study identifies replicable, region-specific grey matter atrophy in the superior frontal cortex and thalamus across two independent RRMS cohorts. It shows that atrophy in these regions occurs earlier and more rapidly in MS than in healthy controls who are approximately 20 years older. Frontal lobe atrophy was most consistently associated with disability progression. Yet, associations between atrophy and changes in clinical scores were weak and sample dependent.

*How this study might affect research, practice or policy:* Our findings suggest to further explore frontal grey matter atrophy as a potential imaging biomarker for MS progression. This could inform future clinical trials, improve prognostic models, and guide the development of personalized treatment strategies based on region-specific brain changes.

## Introduction

Despite significant technological and pharmacological advancements, further research is needed to achieve a deeper understanding of the disease mechanisms and development of reliable biomarkers of multiple sclerosis. Imaging markers bear potential, and for example, grey matter (GM) loss has been suggested as an early prognostic biomarker.^1^ While numerous studies have investigated whole brain GM volume loss in MS, few have examined region-level changes over time.^2–4^ Emerging evidence suggests distinct region-level volumetric differences between people with MS (pwMS) and healthy controls (HC), particularly reductions in subcortical volumes, such as the thalamus, and superior cortical regions such as the cingulate gyrus.^4,5^ Moreover, regions like the thalamus and frontal lobes, have been strongly linked to disability progression, cognitive decline, psychiatric and sexual dysfunctions in pwMS.^2,6–8^ To establish brain volumes in these key regions as biomarkers, further validations and replications are necessary. This is underscored by significant variability in atrophy patterns which can be observed among pwMS.^5,7,9,10^ Additionally, previous studies often lack the statistical power to observe small effects, typically do not utilize longitudinal designs to better approximate causality, and rarely include longitudinal comparisons between pwMS and HC. Finally, replications of specific findings are the exception rather than the norm.

Hence, for clinical translatability, longitudinal case-control comparisons are essential to map regional brain atrophy patterns. Such data provide insight into trajectories rather than mere correlations, allowing for analyses on temporal dynamics. The explanatory value of regionally accelerated atrophy for clinical outcomes could pave the way for the development of new image-derived biomarkers. Furthermore, comparing regional GM atrophy between pwMS and HC, can help determine the region-specific pattern of accelerated brain ageing in MS. However, comparing ageing processes poses challenges, as brain volumes remain relatively stable from the twenties to forties.^11^ Detecting subtle changes is particularly challenging over short scanning intervals, where known measurement variability can exceed the actual rate of brain change.^12^ Since the rate of brain atrophy accelerates with age,^13–17^ using samples from older HC provides a more realistic comparison of ageing processes with younger pwMS.

In this study, we therefore investigated whether generalisable patterns of brain GM volume changes can be identified by replicating our findings in two independent cohorts of pwMS. We studied age-related associations and atrophy patterns in grey matter regions by utilizing a large longitudinal magnetic resonance imaging (MRI) dataset of HC spanning a large age range (see Figure 1). This allowed us to compare atrophy across samples to determine whether RRMS is associated with accelerated atrophy in different brain regions. Finally, we investigated whether these patterns were associated with disability progression, fatigue and cognitive decline.

**Figure 1.**
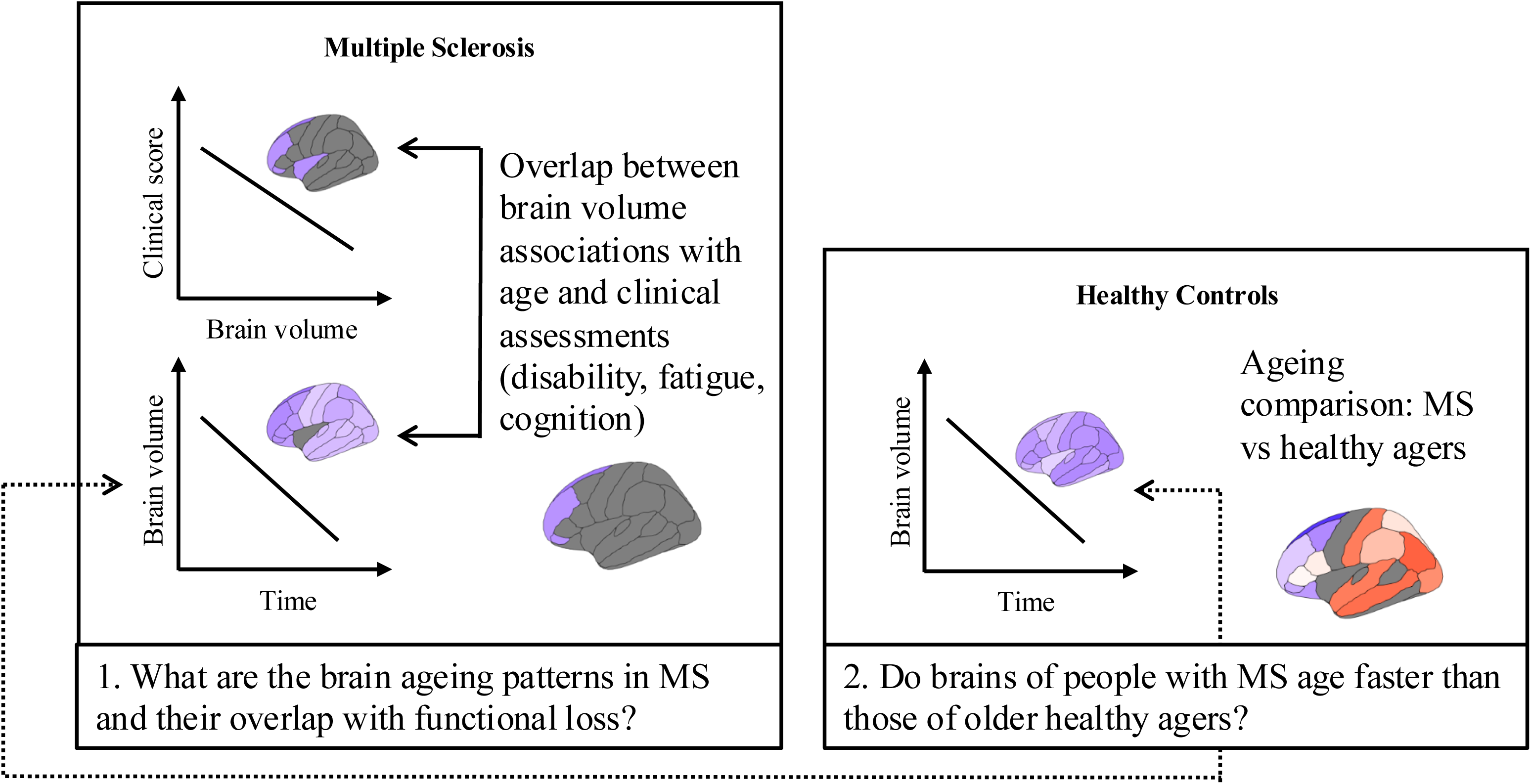
Key research questions and analysis workflow.

## Methods

### Participants

Multiple RRMS and HC datasets were used, which were sample across time (see Figure 2).

**Figure 2.**
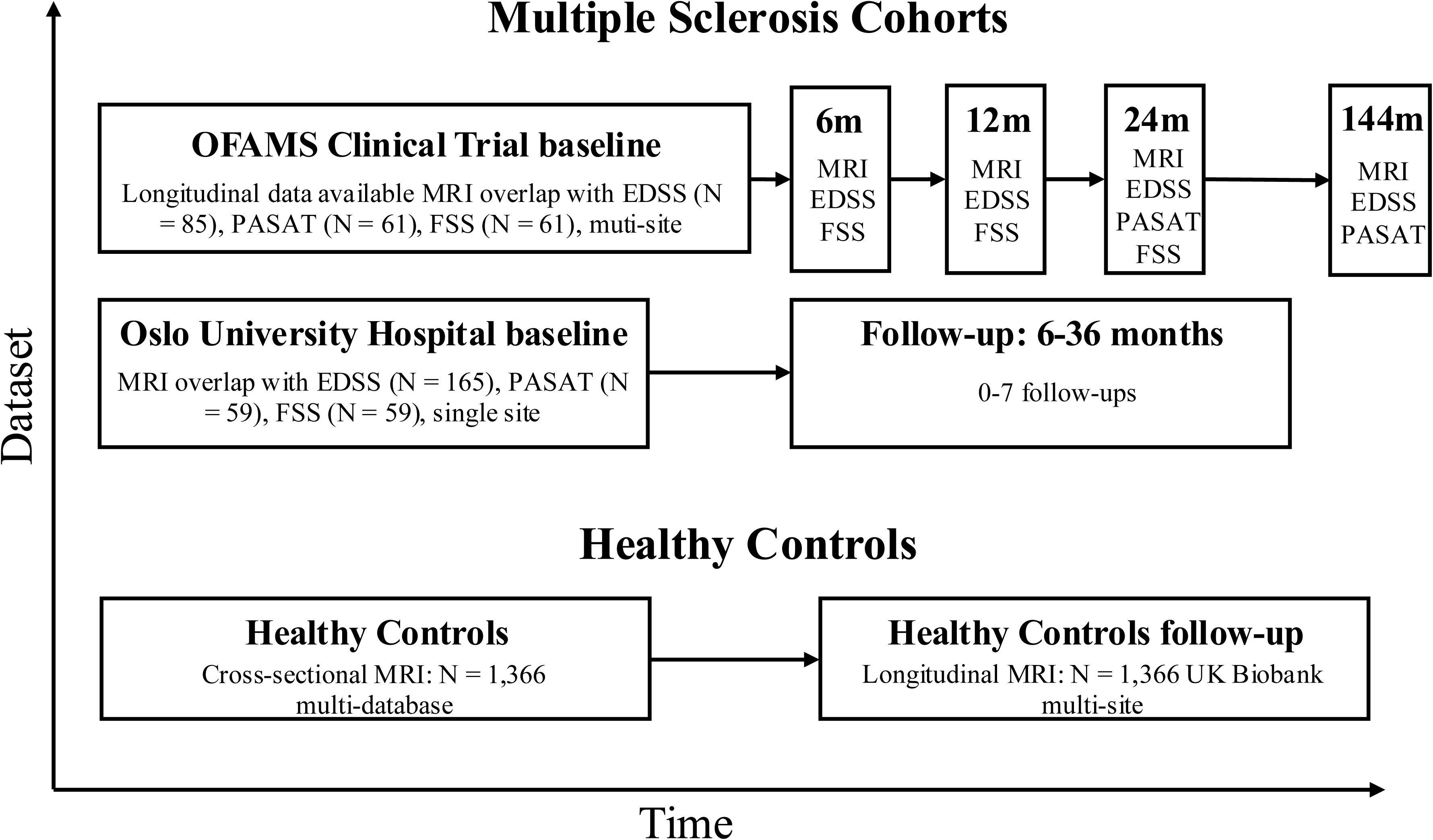
Overview of datasets, sampling and data availability. m = months. *Note*: Data availability indicated in the baseline condition describes the number of participants across time.

The first RRMS dataset included 88 pwMS with RRMS who previously participated in a multicentre clinical trial on omega-3 fatty acid in MS (OFAMS)^18^. The trial entailed data collection every 6 months over 2 years, followed by a single follow-up visit 10 years after the original trial concluded, with N=85 subjects participating in the 10-year follow-up visit (T_1_w-scans=250). The study was approved by the Norwegian Regional Committees for Medical and Health Research Ethics (REK-814351, REK-2016/1906) and registered as clinical trial (clinicaltrials.gov identifier: NCT00360906).

The second RRMS dataset was collected at the Oslo university hospital (OUH)^19,20^ using all available data points with MRI available, being up to 8 follow-ups in a 5-12 month interval. Altogether 302 pwMS with RRMS participated (T_1_w-scans=690; ethics approval: REK-2011/1846).

Both RRMS cohorts included data on disability (Expanded Disability Status Scale, EDSS), cognitive function, using standardized 3 second Paced Auditory Serial Addition Test (PASAT) scores, and fatigue (Fatigue Severity Scale, FSS). EDSS scores were available at all MRI timepoints, however, PASAT scores were only available at baseline, month 24 and at 10-year follow-up for all OFAMS participants (N=85, overlap with T_1_w-scans=282) and across MRI sessions in OUH data (N=53, overlap with T_1_w-scans=82). FSS were obtained at baseline, month 6, 12 and 24 in OFAMS (N=85, overlap with T_1_w-scans=127), and across MRI sessions in OUH data (N=82, T_1_w-scans=125). Missing data resulted from participant no-shows, often related to relapses or other health-related issues, as well as protocol changes implemented during the course of data collection We used the available longitudinal data from the original 48,044 UK Biobank HCs, being N = 2163 with 4,326 scans. After matching the age-range of the two RRMS datasets (N=386, T_1_w-scans=940, age range: 18-67), a final HC datasets of N=1,586 with T_1_w-scans=3,172 was obtained, with a mean follow-up time of 2.44± 0.73 years. Analyses on UK Biobank data were conducted under application 27412 and ethics approval for the UK Biobank was previously obtained from the UK National Research Ethics Service (11/NW/0382). All participants gave their written informed consent. All methods were performed in accordance with the relevant guidelines and regulations (Declaration of Helsinki).

### Magnetic resonance imaging (MRI) and data processing

T_1_-weighted MRI data were obtained using various protocols, scanners, sites, and field strengths (1.5T or 3T). An overview of the acquisition protocols can be found in the original studies and Supplement (HCs: Supplemental Table 1, OFAMS: Supplemental Table 3 and original study^18^, OUH: Supplemental Note 4 and original studies^19,21^). After lesion filling, volumes were extracted using the longitudinal pipeline of FreeSurfer (OFAMS: 7.1.1, OUH: 6.0.0 for OUH data) and averaged for each of the 34 cortical from each hemisphere specified in the Desikan-Killany atlas. Additionally, we used the 6 subcortical grey matter regions provided by the standard FreeSurfer subcortical parcellation, including the thalamus pallidum, amygdala, hippocampus, putamen, caudate, in both hemispheres. Note that version differences were addressed in the next step, during harmonisation.

### MRI Data harmonisation

We applied ComBat, a data harmonisation pipeline, originally developed for batch effects in laboratory samples^22^ The data were harmonised across scanners within both RRMS datasets to allow for replications of the findings from one dataset to another. HC data were harmonised independent from the MS data to avoid spillover harmonisation effects between RRMS and HC datasets. As a quality control, we also report results without using ComBat and instead the inclusion of the scanner as a random intercept in the Supplement.

### Statistical analysis

First, we assessed the rate of volume change in the longitudinal samples using linear mixed models with a random intercept at the level of the individual, associating age with regional brain volume change, controlling for sex and the estimated total intracranial volume. To later compare model coefficients of the random intercept models between HC and MS, the model structure was applied in both groups. We tested for 80 brain regions, leading to an adjusted 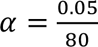, which was applied to each of the tests which attempted to replicate the findings across samples.

Second, we extracted the selected brain regions with significant volume change in step 1, across both RRMS datasets. As previously described,^23^ these coefficients, or slopes, were statistically compared against the coefficients observed in the longitudinal UK Biobank sample (for which longitudinal data were available).

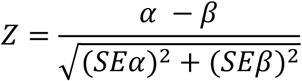

α and β are the respective standardized coefficients and *SE*α and *SE*β representing their standard errors. For display, we present all significant Z-values in Figure 4, and Bonferroni adjusted significant values in-text (marked p_Bonferroni_) for data combining both cohorts with 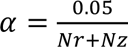, where Nr=80 is the number of brain regions, Nz the number of Z-comparisons, equal to the number of significant replicating age-related volume changes across RRMS samples.

Third, to highlight the clinical relevance of brain volumes, we assessed longitudinal associations between the volume changes and EDSS scores for each dataset individually: OUH and OFAMS data individually, as well as a combined dataset to maximise power, using random intercept models of the same structure as reported in step one but adding EDSS (over time) as explanatory variable of brain volume change across time. Again, we present uncorrected alpha and an adjusted 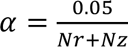. In case of no effect surviving the multiple comparison correction, we present uncorrected p-values, which are not marked as p_Bonferroni_. The full analysis workflow is presented in Figure 1.

For comparability, we report standardized regression coefficients and 95% confidence intervals in square brackets. When reporting multiple effects of the same directionality at once, confidence intervals are not reported. The significance-level was set at a conventional α=0.05, and Bonferroni-adjusted for multiple comparisons. To reduce the convoluting effect of age differences between HC and RRMS samples, the older, longitudinal UK Biobank sample was restricted to participants not exceeding the maximum age of the RRMS samples. Replicability was defined as significant effects of the same direction being observed in both independent samples. For statistical analyses, R version 4.5.0 was used, for spatial representations of the examined brain areas, we used the ggseg R package, and for power simulations the simr package, and complete cases included for individual tests. We used all datasets for which MRI data were available. Simulation-based statistical power estimates for the executed tests was reported in Supplemental Note 3.

## Results

Clinical scores, and in particular PASAT and FSS were relatively stable in both samples, with EDSS showing the largest changes over time (Table 1).

**Table 1.** Overview of the cohorts’ first and last visit with available MRI. Note that PASAT scores were only available for N=54 in the OUH sample.

| Dataset | OFAMS |  | OUH |  | UK Biobank |  |
| --- | --- | --- | --- | --- | --- | --- |
| Assessment | First | Final | First | Final | First | Final |
| Age | 38.9±8.4 | 49.0±9.1 | 38.6±9.9 | 39.5±9.5 | 59.1±5.2 | 61.0±5.0 |
| Females | 54 | 50 | 220 | 62 | 1207 | 1207 |
| Males | 30 | 27 | 82 | 12 | 956 | 956 |
| EDSS | 1.9±0.8 | 2.6±1.4 | 2.0±1.2 | 2.1±1.5 |  |  |
| PASAT | 47.8±9.4 | 45.7±12.2 | 48.3±9.0 | 48.0±8.5 |  |  |
| FSS | 4.5±1.6 | 4.4±1.8 | 4.0±1.9 | 4.0±2.0 |  |  |

When examining brain GM volume change in pwMS in the two RRMS cohorts (Table 1), we found replicable patterns of grey matter loss in multiple cortical and subcortical regions (Figure 3, Supplemental Table 1). The strongest replicable effect of age on grey matter volume were found in both hemispheres of the *SFC* (OUH: β_age_<-0.27, p_Bonferroni_<0.001; OFAMS: β_age_<-0.51, p_Bonferroni_<0.001), *pars orbitalis* (OUH: β_age_<-0.25, p_Bonferroni_<0.001; OFAMS: β_age_<-0.41, p_Bonferroni_<0.001), and *thalami* (OUH: β_age_<-0.20, p_Bonferroni_<0.001; OFAMS: β_age_<-0.52, p_Bonferroni_<0.001) in each hemisphere. The overall effect on whole brain GM atrophy was weaker than these largest observed regional effects in OUH (β_age_=-0.25 [-0.15, -0.35], p_Bonferroni_<0.001), and OFAMs data (β_age_=-0.36 [-0.28, -0.45], p_Bonferroni_<0.001).

**Figure 3.**
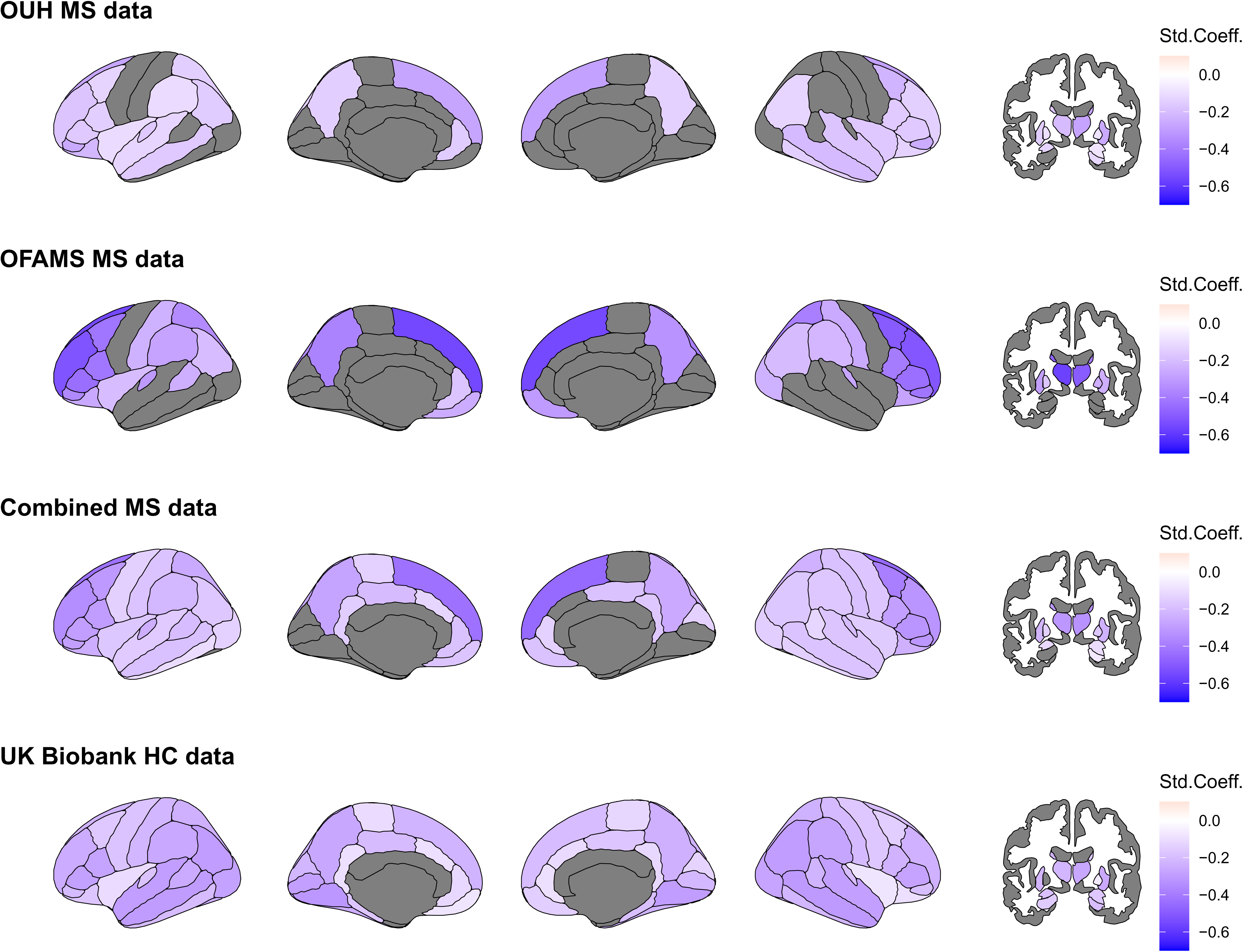
Volume change in RRMS and the UK Biobank. Top row: ageing effects in the OUH sample. Second row: ageing effects in the OFAMS sample. Third row: ageing effects in both RRMS cohorts combined. Fourth row: ageing effects in the longitudinal UK Biobank sample of healthy controls. The two left columns represent the left cortex, followed by two columns representing the right cortex, and the rightmost column of a coronal view for subcortical changes. Grey regions indicate non-significant age effects (Bonferroni-corrected p≥0.05), blue areas indicate significant age associations after Bonferroni correction, with darker colours indicating stronger relationship indicated here by standardized coefficients (Std.Coeff.). For comparison, the effect of age on regional brain volumes in older HCs in the longitudinal sample (UK Biobank) was smaller than in pwMS (Supplemental Note 5).

When statistically comparing the standardized longitudinal effects (slopes) of age on brain volumes between pwMS and a 20 year older cohort of HCs (Figure 4), we found replicable faster GM atrophy in pwMS in the right SFC (OUH: Z=-2.41, p = 0.008; OFAMS: Z=-6.49, p<0.001), the right caudal middle frontal cortex (OUH: Z=-2.09, p = 0.018; OFAMS: Z=-6.38, p<0.001), right caudate (OUH: Z=-2.46. p = 0.007; OFAMS: Z=-2.10, p = 0.018), and left frontal pole (OUH: Z=-2.43. p = 0.007; OFAMS: Z=-2.99, p = 0.001). For the other regions, atrophy rates were either inconsistently different from HC across the two MS datasets or non-significant, indicating no difference in ageing slopes between older HC and pwMS. For a full overview of differences in effects see Supplemental Table 2.

**Figure 4.**
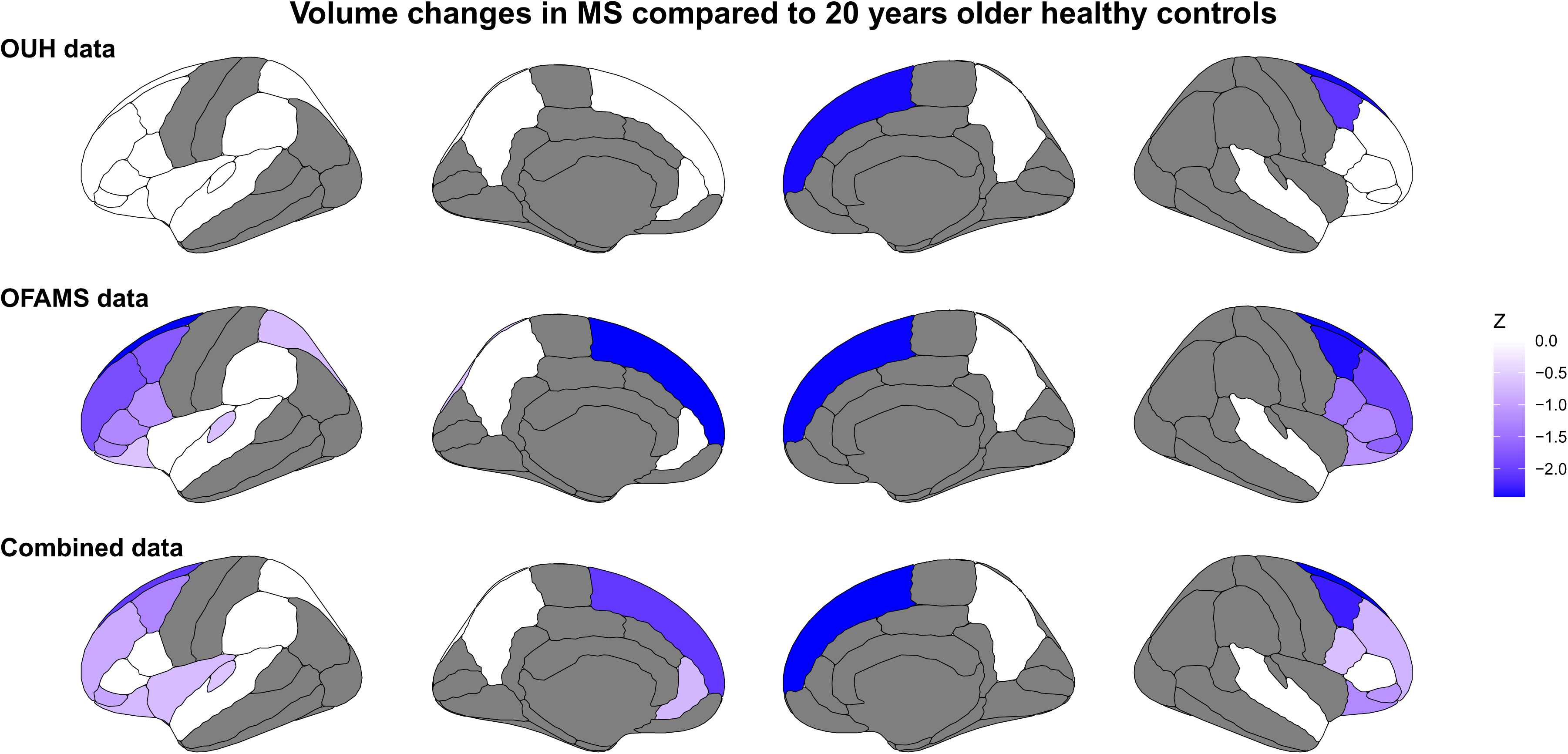
Volume changes in younger pwMS compared to 20 years older healthy controls. The two left columns represent the left cortex, and the two right columns represent the right cortex. The regions considered for this analysis were selected based on cross-sample replicated significant brain changes/atrophy. Blue colours indicate faster volume change in RRMS, white colour indicates no difference (p>0.05). The top two rows indicate comparisons between standardised regression coefficients of single RRMS datasets and UK Biobank data. The bottom row indicates such comparisons between coefficients from combined RRMS data and UK Biobank healthy controls. The presented significance level is not adjusted for multiple comparisons. Only cortical areas are displayed due to the lack of significant subcortical areas, except the caudate and thalamus, which are highlighted in-text.

None of the significant regional atrophy was significantly related to PASAT, EDSS or FSS in both samples. At an uncorrected threshold of 0.05, across samples, changes in EDSS were significantly related to atrophy in the right (OFAMS: β_EDSS_ = -0.11 [-0.02, -0.21], p = 0.026, OUH: β_EDSS_ = -0.10 [-0.05, -0.14], p<0.001), and left thalamus (OFAMS: β_EDSS_ = -0.13 [-0.03, -0.24], p = 0.01, OUH: β_EDSS_ = -0.09 [-0.05, -0.13], p < 0.001), right pallidum (OFAMS: β_EDSS_ = -0.13 [-0.01, -0.25], p = 0.02, OUH: β_EDSS_ = -0.07 [-0.01, -0.13], p = 0.03), left hippocampus (OFAMS: β_EDSS_ = -0.19 [-0.07, -0.31], p = 0.002, OUH: β_EDSS_ = -0.11 [-0.06, -0.15], p < 0.001), and left entorhinal cortex (OFAMS: β_EDSS_ = -0.20 [-0.08, -0.32], p = 0.001, OUH: β_EDSS_ = -0.05 [-0.01, -0.09], p = 0.03). Whole brain atrophy was stronger associated with changes in EDSS in both samples (OFAMS: β_EDSS_= -0.33 [-0.44, -0.23], p<0.001, OUH: β_EDSS_= -0.20 [-0.25, -0.16], p<0.001).

Such associations were not found for FSS (OFAMS: β_fatigue_=-0.13 [-0.02, -0.24] p = 0.018, OUH: p = 0.331), or PASAT (OFAMS: p = 0.327, OUH: β_PASAT_=0.08 [0.009, 0.15], p = 0.030). For an analysis approach combining the two RRMS cohorts see Supplemental Note 1. The presented ageing-effects were reproduced when using scanner as a fixed effect instead of ComBat harmonisation (Supplemental Figures 1-2). However, the smaller effects of clinical assessments on brain volumes could not be reproduced in both examined RRMS cohorts without appropriate harmonisation (Supplemental Note 2).

## Discussion

We used two independent RRMS cohorts to replicate regional GM atrophy patterns, examine their associations with clinical outcomes, and compare these patterns with those observed during normal aging. Consistent with previous studies,^5,7,9,10^ our findings highlight atrophy in the SFC and thalamus, which in this study is outlined as RRMS-specific, unique ageing process. This contrasts a more wide-spread and less significant pattern of healthy and slower ageing in approximately 20 years older HCs in SFC and thalamus. Additionally, atrophy effects were better captured on the regional than whole-brain level. This suggests that there are generally faster atrophy rates in MS than in HC and that this atrophy is region-specific. Clinical outcomes were relatively stable, which might be a reason for the non-significant associations between atrophy and changes in FSS and PASAT after corrections for multiple testing. Nevertheless, an uncorrected significance level suggest that thalamus atrophy is associated with EDSS changes, highlighting the thalamus as a clinically relevant MS-specific and potentially vulnerable brain region. At the same time, associations between whole-brain volumetric and EDSS changes were stronger than region-level level changes.

Our findings align with previous cross-sectional and longitudinal studies on MS emphasizing frontal GM atrophy,^5,7,24^ which has been associated with a range of MS-related disabilities, including psychiatric signs, cognitive decline, autonomic and sexual dysfunctions.^6–8^ However, there is variability across findings concerning the exact atrophy pattern, with different studies highlighting also other areas than presented here (strongest associations: frontal lobes, thalami and pars orbitalis). For example, in line with our findings, one study shows the strongest atrophy pattern in the SFC, yet also in the insula and paracentral gyrus.^24^ Similarly, while other studies also report the frontal lobe as one of the most significantly changing regions, strong atrophy patterns were also found in the cingulate,^5^ or parietal and occipital gyri.^7^ In contrast to our findings, which present replicable associations between EDSS and volume changes in thalami, pallidum, hippocampus, and entorhinal cortex, another longitudinal study highlights praecuneus, insula, parahippocampal gyrus and cingulate instead.^25^ Additionally, several recent studies presented an association between thalamic volume changes and EDSS changes.^2,25,26^ Yet, our data suggested smaller associations between volumes and clinical outcomes. A possible explanation might be that most of the pwMS in our cohorts were only very slowly progressing, and follow-up times in the OUH data were relatively short. This is concordant with the progression of most pwMS on modern treatment plans and especially early diagnosis, underlining the usefulness of the presented findings.

Moreover, our findings highlight the differential vulnerability of distinct brain regions to RRMS-related neurodegeneration, each contributing uniquely and interactively to the overall symptomatology.^2,6–8^ While regional atrophy provides valuable insights, it is the integration of these regions, representing complex network-level dysfunction, that likely underlies the heterogeneous clinical presentation of MS.^27–30^ This interdependence poses a significant challenge for research, as focusing solely on region-specific effects may overlook broader patterns of distributed dysfunction. Nonetheless, the spatial detail provided by a region-based approach makes it superior to analyses of whole-brain volumes for understanding the nuanced impacts of MS. Part of the diverse set of symptoms which characterise MS may be explained by the high level of connectivity between the frontal lobes and other brain regions. Functional connectivity MRI studies on pwMS have consistently demonstrated the involvement of the frontal lobes (see for review^31^). Another key region identified in this study was the thalamus, which holds prognostic value due to its susceptibility to atrophy processes early in the disease course and association with physical disability.^2,7,8,10,32^ Ultimately, MS may be best understood as a network of behavioural and biomarkers, encompassing not only regional atrophy but also a constellation of image-derived markers and functional disruptions.^27–30^ In this context, our study provides a foundational step by identifying generalizable patterns of brain atrophy, which can serve as a basis for more detailed investigations into the network-level mechanisms underlying disease progression.

The present study has several notable strengths compared to previous studies. First, we leveraged two independent, longitudinal RRMS cohorts, allowing for replication of findings across different samples and timescales. Replications across samples provide a crucial validation step, allowing specifically to better account for measurement error than when examining a single isolated sample. Additionally, difference between samples, the extended follow-up period in the OFAMS cohort (up to 12 years) and the shorter but more recent OUH dataset, provided complementary insights into disease progression over varying temporal scales. Second, the inclusion of HC allowed us to benchmark MS-specific atrophy patterns against the trajectories of normal aging. Third, we harmonised the imaging data to minimize technical variability and enhance comparability of findings. Fourth, most previous studies do not report standardized effects, making comparability across studies difficult. Standardized effects, as presented here, provide a useful tool for the comparability of study results. Finally, our region-specific approach enabled us to identify key neuroanatomical targets, such as the SFC and thalamus, that are linked to clinical outcomes, establishing a foundation for future research into MS-related neurodegeneration.

There are limitations to our study, primarily related to sample-specific variability origination from multiple sources of error, and lack of disability progression. While we identified distinct and replicable atrophy patterns, these sources of error may have limited the discovery rate of true effects across samples. Data from the OFAMS trial were sampled over a longer time (12 years) compared to those from the OUH sample (2 years), potentially explaining the stronger effects of age observed in the OFAMS sample. Measurement errors in the OUH sample may have been larger than the percentage change observed during the shorter sampling period.^11,12^ The OUH data were collected at a single hospital, while OFAMS data were collected at multiple sites with different scanners and protocols, introducing variability. Differences in available disease-modifying treatments in the OFAMS study and the OUH data may have influenced the results, as data collection started in 2004 and 2012, respectively.

For practical reasons, we used two different versions of FreeSurfer (6.0.0 and 7.1.1) for data processing. However, the variability due to these processing choices is expected to be minor.^33,34^ The Desikan-Killiany atlas-based regional averages used in this study are less susceptible to spatial inaccuracies compared to atlases with smaller parcels or voxel-level analyses. Even for FreeSurfer versions further apart (5.3.0 and 7.4.1) than reported here (6.0.0 and 7.1.1), the correlations between features are shown to be acceptable (mean r = 0.88).^35^ These parcels do, however, have reduced spatial specificity, both for detecting true underlying effects and for identifying version-induced variability in the spatial pattern.

Finally, while a genuine part of the presented data, the slow disability progression might be a reason for small associations between changes in brain volumes and clinical outcomes. While the relative stability in PASAT scores might originate from practice effects,^36^ consistent with the observations in our cohorts, fatigue measures have previously been reported to be relatively stable over time.^37^ Moreover, non-significant associations between measures of fatigue and MRI have previously been reported.^38,39^ and the brain structural underpinnings of fatigue In multicentre studies, harmonisation is crucial to mitigate technical, non-biological effects. For this study, we applied the longComBat pipeline specifically designed for longitudinal data.^40^ While the ComBat approach is considered state-of-the-art and has been shown to effectively remove scanner effects,^40^ it cannot completely be excluded that the method also inadvertently alters observable biological effects. While we found some differences between harmonisation strategies (ComBat vs fixed effects regression) and no harmonisation, these differences remained relatively small. However, whether their origin is biological or non-biological is unclear. To control for multiple comparisons, we applied a conservative Bonferroni-correction for α-level adjustment. While no definite guideline exists on which correction method is optimal, this conservative approach may have led to false negatives for detecting small effects, particularly in associations between clinical outcomes and GM atrophy. Given restricted statistical power to detect subtle associations, variability across participants, and relatively stable clinical outcome measures, the reported brain-behaviour findings should be investigated in larger and more densely sampled cohorts.

In conclusion, we observed accelerated GM degeneration the frontal lobes and thalami in RRMS. This atrophy progressed more rapidly than in 20-year older HC and, for thalami, was associated with disability-progression. Our findings suggest that accelerated atrophy in RRMS occurs decades earlier than compared to HC. Regional atrophy in frontal lobes and thalami might be regions of interest to develop disease imaging markers reflecting RRMS disease courses.

## Open Data and Materials

Summary statistics and utilized code can be found in the GitHub repository: https://github.com/MaxKorbmacher/MSatrophy. Multiple of the utilized dataset are sensitive, require IRB approval for usage, and can therefore not be openly shared.

## Data Availability

Summary statistics (anonymous) and utilized code can be found in the GitHub repository: https://github.com/MaxKorbmacher/MSatrophy. Multiple of the utilized dataset are sensitive, require IRB approval for usage, and can therefore not be openly shared.

## Acknowledgments

We extend our sincere gratitude to the people with MS who participated in this study, whose contributions are invaluable. This research originated from different research efforts: one national intervention study (OFAMS) and ten years follow-up examinations, as well the ongoing data collections at the MS research group at the Oslo University Hospital.

We thank the OFAMS study group at numerous hospitals across Norway, as well as the group of clinicians and researchers at the Oslo University Hospital.

Finally, we want to extend our gratitude to the thousands of scanned participants in the UK Biobank study and facilitators who made the study possible.

## Competing interests

OAA has received a speaker’s honorarium from Lundbeck, Janssen, Otsuka and Lilly, and is a consultant to Coretechs.ai and Precision Health.

LTW is a minor shareholder of baba.vision.

KMM has served on scientific advisory board for Alexion, received speaker honoraria from Biogen, Lundbeck, Novartis and Roche, and has participated in clinical trials organized by Biogen, Merck, Novartis, Otivio, Roche and Sanofi.

EAH received honoraria for advisory board activity from Sanofi-Genzyme, and his department has received honoraria for lecturing from Biogen and Merck.

ØT received speaker honoraria from and served on scientific advisory boards of Biogen, Sanofi-Aventis, Merck, and Novartis, and has participated in clinical trials organized by Merck, Novartis, Roche and Sanofi.

SW received speaker honoraria from Biogen, Sanofi-Aventis, and Janssen, and has participated in commissioned research projects funded by Merck, Novartis, and EMD Serono.

MK is a postdoctoral researcher whose career progression depends in part on publishing scientific research. As such, he has a professional interest in disseminating the findings of this study.

The remaining authors declare no other competing interests.

## Contributions

MK: Conceptualisation and design of the study, Drafting the manuscript, Statistical analyses. Responsible for the overall content.

All authors contributed in different capacities to data acquisition, administration, or interpretation, as well as critical revisions of the article. All authors approved the final version of the manuscript.

## References

1. Stankoff, B. & Louapre, C. Can we use regional grey matter atrophy sequence to stage neurodegeneration in multiple sclerosis? Brain 141, 1580–1583 (2018).

2. Cagol, A. et al. Association of Brain Atrophy With Disease Progression Independent of Relapse Activity in Patients With Relapsing Multiple Sclerosis. JAMA Neurol. 79, 682– 692 (2022).

3. Nygaard, G. O. et al. Cortical thickness and surface area relate to specific symptoms in early relapsing–remitting multiple sclerosis. Mult. Scler. J. 21, 402–414 (2015).

4. Lansley, J., Mataix-Cols, D., Grau, M., Radua, J. & Sastre-Garriga, J. Localized grey matter atrophy in multiple sclerosis: A meta-analysis of voxel-based morphometry studies and associations with functional disability. Neurosci. Biobehav. Rev. 37, 819–830 (2013).

5. Eshaghi, A. et al. Progression of regional grey matter atrophy in multiple sclerosis. Brain 141, 1665–1677 (2018).

6. Andravizou, A. et al. Brain atrophy in multiple sclerosis: mechanisms, clinical relevance and treatment options. Autoimmun. Highlights 10, 7 (2019).

7. Eijlers, A. J. C. et al. Cortical atrophy accelerates as cognitive decline worsens in multiple sclerosis. Neurology 93, e1348–e1359 (2019).

8. Grey matter pathology in multiple sclerosis - The Lancet Neurology. https://www.thelancet.com/journals/lancet/article/PIIS1474-4422(08)70191-1/abstract.

9. Martinez-Heras, E. et al. Diffusion-based structural connectivity patterns of multiple sclerosis phenotypes. J. Neurol. Neurosurg. Psychiatry 94, 916–923 (2023).

10. Meijboom, R. et al. Patterns of brain atrophy in recently-diagnosed relapsing-remitting multiple sclerosis. PLOS ONE 18, e0288967 (2023).

11. Wang, M.-Y. et al. The within-subject stability of cortical thickness, surface area, and brain volumes across one year. 2024.06.01.596956 Preprint at 10.1101/2024.06.01.596956 (2024).

12. Vidal-Piñeiro, D. et al. Reliability of structural brain change in cognitively healthy adult samples. Imaging Neurosci. (2025) doi:10.1162/imag_a_00547.

13. Korbmacher, M. et al. Distinct longitudinal brain white matter microstructure changes and associated polygenic risk of common psychiatric disorders and Alzheimer’s disease in the UK Biobank. Biol. Psychiatry Glob. Open Sci. 100323 (2024) doi:10.1016/j.bpsgos.2024.100323.

14. Fujita, S. et al. Characterization of Brain Volume Changes in Aging Individuals With Normal Cognition Using Serial Magnetic Resonance Imaging. *JAMA Netw*. Open 6, e2318153 (2023).

15. Bethlehem, R. A. et al. Brain charts for the human lifespan. Nat 604, 525–533 (2022).

16. Korbmacher, M. et al. Brain-wide associations between white matter and age highlight the role of fornix microstructure in brain ageing. HBM 44, (2023).

17. Korbmacher, M. et al. Brain asymmetries from mid-to late life and hemispheric brain age. Nat. Commun. 15, 956 (2024).

18. Torkildsen, O. et al. ω-3 fatty acid treatment in multiple sclerosis (OFAMS Study): a randomized, double-blind, placebo-controlled trial. Arch. Neurol. 69, 1044–1051 (2012).

19. Høgestøl, E. A. et al. Cross-Sectional and Longitudinal MRI Brain Scans Reveal Accelerated Brain Aging in Multiple Sclerosis. Front. Neurol. 10, 450 (2019).

20. Sowa, P. et al. Restriction spectrum imaging of white matter and its relation to neurological disability in multiple sclerosis. Mult. Scler. Houndmills Basingstoke Engl. 25, 687–698 (2019).

21. Sowa, P. et al. Restriction spectrum imaging of white matter and its relation to neurological disability in multiple sclerosis. Mult. Scler. Houndmills Basingstoke Engl. 25, 687–698 (2019).

22. Johnson, W. E., Li, C. & Rabinovic, A. Adjusting batch effects in microarray expression data using empirical Bayes methods. Biostatistics 8, 118–127 (2007).

23. Clogg, C. C., Petkova, E. & Haritou, A. Statistical methods for comparing regression coefficients between models. Am. J. Sociol. 100, 1261–1293 (1995).

24. Steenwijk, M. D. et al. Cortical atrophy patterns in multiple sclerosis are non-random and clinically relevant. Brain 139, 115–126 (2016).

25. Ziccardi, S. et al. Early regional cerebral grey matter damage predicts long-term cognitive impairment phenotypes in multiple sclerosis: a 20-year study. Brain Commun. 6, fcae355 (2024).

26. Magon, S. et al. Volume loss in the deep gray matter and thalamic subnuclei: a longitudinal study on disability progression in multiple sclerosis. J. Neurol. 267, 1536– 1546 (2020).

27. Eijlers, A. J. C. et al. Increased default-mode network centrality in cognitively impaired multiple sclerosis patients. Neurology 88, 952–960 (2017).

28. Kennedy, K. E. et al. Multiscale networks in multiple sclerosis. PLOS Comput. Biol. 20, e1010980 (2024).

29. Fleischer, V. et al. Prognostic value of single-subject grey matter networks in early multiple sclerosis. Brain 147, 135–146 (2024).

30. Martí-Juan, G. et al. Using The Virtual Brain to study the relationship between structural and functional connectivity in patients with multiple sclerosis: a multicenter study. Cereb. Cortex 33, 7322–7334 (2023).

31. Rocca, M. A., Schoonheim, M. M., Valsasina, P., Geurts, J. J. G. & Filippi, M. Task- and resting-state fMRI studies in multiple sclerosis: From regions to systems and time-varying analysis. Current status and future perspective. NeuroImage Clin. 35, 103076 (2022).

32. Schoonheim, M. M. et al. Thalamus structure and function determine severity of cognitive impairment in multiple sclerosis. Neurology 84, 776–783 (2015).

33. Sokołowski, A. et al. The impact of FreeSurfer versions on structural neuroimaging analyses of Parkinson’s disease. bioRxiv 2024–11 (2024).

34. Hedges, E. P. et al. Reliability of structural MRI measurements: The effects of scan session, head tilt, inter-scan interval, acquisition sequence, FreeSurfer version and processing stream. NeuroImage 246, 118751 (2022).

35. Korbmacher, M., Westlye, L. T. & Maximov, I. I. FreeSurfer version-shuffling can enhance brain age predictions. NeuroImage Rep. 4, 100214 (2024).

36. Castrogiovanni, N. et al. Longitudinal Changes in Cognitive Test Scores in Patients With Relapsing-Remitting Multiple Sclerosis: An Analysis of the DECIDE Dataset. Neurology 101, e1–e11 (2023).

37. Téllez, N. et al. Fatigue in multiple sclerosis persists over time: a longitudinal study. J. Neurol. 253, 1466–1470 (2006).

38. Kluger, B. M., Krupp, L. B. & Enoka, R. M. Fatigue and fatigability in neurologic illnesses. Neurology 80, 409–416 (2013).

39. Meijboom, R. et al. Fatigue in early multiple sclerosis: MRI metrics of neuroinflammation, relapse and neurodegeneration. Brain Commun. 6, fcae278 (2024).

40. Beer, J. C. et al. Longitudinal ComBat: A method for harmonizing longitudinal multi-scanner imaging data. NeuroImage 220, 117129 (2020).

