## Supplement for "Accelerated grey matter degeneration in relapsing remitting multiple sclerosis"

### Supplemental Tables

***Supplemental Table 1*. Longitudinal standardized associations of age with brain volumes, controlling for sex, and total intracranial volume across samples.**

| ROI | Beta | $CI_{high}$ | $CI_{low}$ | p | $p_{Bonf}$ | data |
|---|---|---|---|---|---|---|
| lh_bankssts_volume | -0.06 | 0 | -0.13 | 0.05 | 1 | OUH |
| lh_caudalanteriorcingulate_volume | -0.08 | -0.03 | -0.13 | 0 | 0.21 | OUH |
| lh_caudalmiddlefrontal_volume | -0.13 | -0.06 | -0.2 | 0 | 0.05 | OUH |
| lh_cuneus_volume | 0.06 | 0.14 | -0.01 | 0.1 | 1 | OUH |
| lh_entorhinal_volume | 0.08 | 0.15 | 0 | 0.05 | 1 | OUH |
| lh_fusiform_volume | -0.04 | 0.03 | -0.1 | 0.25 | 1 | OUH |
| lh_inferiorparietal_volume | -0.15 | -0.09 | -0.21 | 0 | 0 | OUH |
| lh_inferiortemporal_volume | -0.06 | 0 | -0.12 | 0.05 | 1 | OUH |
| lh_isthmuscingulate_volume | -0.02 | 0.04 | -0.08 | 0.47 | 1 | OUH |
| lh_lateraloccipital_volume | -0.04 | 0.04 | -0.11 | 0.33 | 1 | OUH |
| lh_lateralorbitofrontal_volume | -0.21 | -0.15 | -0.28 | 0 | 0 | OUH |
| lh_lingual_volume | 0.07 | 0.14 | 0 | 0.05 | 1 | OUH |
| lh_medialorbitofrontal_volume | -0.07 | 0 | -0.14 | 0.06 | 1 | OUH |
| lh_middletemporal_volume | -0.16 | -0.1 | -0.22 | 0 | 0 | OUH |
| lh_parahippocampal_volume | 0.06 | 0.14 | -0.01 | 0.08 | 1 | OUH |
| lh_paracentral_volume | -0.06 | 0.01 | -0.14 | 0.1 | 1 | OUH |
| lh_parsopercularis_volume | -0.14 | -0.08 | -0.21 | 0 | 0 | OUH |
| lh_parsorbitalis_volume | -0.26 | -0.18 | -0.33 | 0 | 0 | OUH |
| lh_parstriangularis_volume | -0.16 | -0.09 | -0.23 | 0 | 0 | OUH |
| lh_pericalcarine_volume | 0.06 | 0.15 | -0.03 | 0.16 | 1 | OUH |
| lh_postcentral_volume | -0.09 | -0.02 | -0.16 | 0.01 | 0.82 | OUH |
| lh_posteriorcingulate_volume | -0.09 | -0.02 | -0.15 | 0.01 | 1 | OUH |

| | | | | | | |
|---|---|---|---|---|---|---|
| lh_precentral_volume | -0.12 | -0.05 | -0.19 | 0 | 0.07 | OUH |
| lh_precuneus_volume | -0.14 | -0.08 | -0.2 | 0 | 0 | OUH |
| lh_rostralanteriorcingulate_volume | -0.15 | -0.1 | -0.21 | 0 | 0 | OUH |
| lh_rostralmiddlefrontal_volume | -0.18 | -0.11 | -0.24 | 0 | 0 | OUH |
| lh_superiorfrontal_volume | -0.27 | -0.21 | -0.33 | 0 | 0 | OUH |
| lh_superiorparietal_volume | -0.14 | -0.08 | -0.21 | 0 | 0 | OUH |
| lh_superiortemporal_volume | -0.12 | -0.06 | -0.19 | 0 | 0.01 | OUH |
| lh_supramarginal_volume | -0.11 | -0.05 | -0.16 | 0 | 0.02 | OUH |
| lh_frontalpole_volume | -0.17 | -0.08 | -0.27 | 0 | 0.03 | OUH |
| lh_temporalpole_volume | 0.07 | 0.15 | -0.02 | 0.14 | 1 | OUH |
| lh_transversetemporal_volume | -0.16 | -0.09 | -0.23 | 0 | 0 | OUH |
| lh_insula_volume | -0.1 | -0.05 | -0.16 | 0 | 0.04 | OUH |
| rh_bankssts_volume | -0.11 | -0.04 | -0.17 | 0 | 0.08 | OUH |
| rh_caudalanteriorcingulate_volume | 0 | 0.06 | -0.06 | 0.94 | 1 | OUH |
| rh_caudalmiddlefrontal_volume | -0.22 | -0.16 | -0.29 | 0 | 0 | OUH |
| rh_cuneus_volume | -0.07 | -0.01 | -0.13 | 0.03 | 1 | OUH |
| rh_entorhinal_volume | 0.1 | 0.17 | 0.02 | 0.01 | 1 | OUH |
| rh_fusiform_volume | -0.05 | 0.01 | -0.11 | 0.12 | 1 | OUH |
| rh_inferiorparietal_volume | -0.12 | -0.06 | -0.19 | 0 | 0.01 | OUH |
| rh_inferiortemporal_volume | -0.14 | -0.08 | -0.2 | 0 | 0 | OUH |
| rh_isthmuscingulate_volume | -0.02 | 0.04 | -0.09 | 0.54 | 1 | OUH |
| rh_lateraloccipital_volume | -0.05 | 0.02 | -0.11 | 0.17 | 1 | OUH |
| rh_lateralorbitofrontal_volume | -0.13 | -0.07 | -0.2 | 0 | 0.01 | OUH |
| rh_lingual_volume | 0 | 0.07 | -0.06 | 0.88 | 1 | OUH |
| rh_medialorbitofrontal_volume | -0.05 | 0.01 | -0.12 | 0.11 | 1 | OUH |
| rh_middletemporal_volume | -0.21 | -0.15 | -0.27 | 0 | 0 | OUH |
| rh_parahippocampal_volume | 0.1 | 0.17 | 0.02 | 0.01 | 1 | OUH |
| rh_paracentral_volume | -0.04 | 0.04 | -0.12 | 0.35 | 1 | OUH |
| rh_parsopercularis_volume | -0.16 | -0.1 | -0.22 | 0 | 0 | OUH |
| rh_parsorbitalis_volume | -0.25 | -0.17 | -0.33 | 0 | 0 | OUH |
| rh_parstriangularis_volume | -0.16 | -0.1 | -0.22 | 0 | 0 | OUH |
| rh_pericalcarine_volume | 0.05 | 0.14 | -0.03 | 0.2 | 1 | OUH |
| rh_postcentral_volume | -0.09 | -0.02 | -0.16 | 0.01 | 1 | OUH |
| rh_posteriorcingulate_volume | -0.06 | 0.02 | -0.13 | 0.13 | 1 | OUH |
| rh_precentral_volume | -0.09 | -0.02 | -0.16 | 0.01 | 0.75 | OUH |
| rh_precuneus_volume | -0.14 | -0.07 | -0.2 | 0 | 0 | OUH |

| | | | | | | |
|---|---|---|---|---|---|---|
| rh_rostralanteriorcingulate_volume | -0.04 | 0.02 | -0.1 | 0.23 | 1 | OUH |
| rh_rostralmiddlefrontal_volume | -0.14 | -0.07 | -0.21 | 0 | 0 | OUH |
| rh_superiorfrontal_volume | -0.28 | -0.22 | -0.35 | 0 | 0 | OUH |
| rh_superiorparietal_volume | -0.1 | -0.03 | -0.17 | 0 | 0.44 | OUH |
| rh_superiortemporal_volume | -0.17 | -0.11 | -0.24 | 0 | 0 | OUH |
| rh_supramarginal_volume | -0.09 | -0.03 | -0.15 | 0 | 0.24 | OUH |
| rh_frontalpole_volume | -0.15 | -0.06 | -0.24 | 0 | 0.08 | OUH |
| rh_temporalpole_volume | 0.1 | 0.18 | 0.01 | 0.02 | 1 | OUH |
| rh_transversetemporal_volume | -0.09 | -0.02 | -0.17 | 0.02 | 1 | OUH |
| rh_insula_volume | -0.17 | -0.11 | -0.22 | 0 | 0 | OUH |
| Left.Amygdala | -0.13 | -0.06 | -0.21 | 0 | 0.06 | OUH |
| Left.Caudate | -0.16 | -0.09 | -0.23 | 0 | 0 | OUH |
| Left.Cerebellum.Cortex | -0.11 | -0.04 | -0.18 | 0 | 0.21 | OUH |
| Left.Hippocampus | -0.07 | 0.01 | -0.15 | 0.07 | 1 | OUH |
| Left.Pallidum | -0.09 | -0.01 | -0.17 | 0.02 | 1 | OUH |
| Left.Putamen | -0.15 | -0.07 | -0.22 | 0 | 0.01 | OUH |
| Left.Thalamus | -0.2 | -0.14 | -0.27 | 0 | 0 | OUH |
| Right.Amygdala | -0.08 | 0 | -0.16 | 0.05 | 1 | OUH |
| Right.Caudate | -0.27 | -0.2 | -0.34 | 0 | 0 | OUH |
| Right.Cerebellum.Cortex | -0.19 | -0.13 | -0.26 | 0 | 0 | OUH |
| Right.Hippocampus | -0.12 | -0.04 | -0.19 | 0 | 0.18 | OUH |
| Right.Pallidum | -0.08 | 0 | -0.17 | 0.04 | 1 | OUH |
| Right.Putamen | -0.26 | -0.18 | -0.33 | 0 | 0 | OUH |
| Right.Thalamus | -0.28 | -0.21 | -0.34 | 0 | 0 | OUH |
| lh_bankssts_volume | -0.29 | -0.19 | -0.38 | 0 | 0 | OFAMS |
| lh_caudalanteriorcingulate_volume | -0.12 | -0.03 | -0.2 | 0.01 | 0.52 | OFAMS |
| lh_caudalmiddlefrontal_volume | -0.43 | -0.32 | -0.53 | 0 | 0 | OFAMS |
| lh_cuneus_volume | -0.12 | -0.01 | -0.23 | 0.03 | 1 | OFAMS |
| lh_entorhinal_volume | 0.07 | 0.2 | -0.07 | 0.33 | 1 | OFAMS |
| lh_fusiform_volume | 0.05 | 0.19 | -0.08 | 0.42 | 1 | OFAMS |
| lh_inferiorparietal_volume | -0.19 | -0.07 | -0.3 | 0 | 0.11 | OFAMS |
| lh_inferiortemporal_volume | -0.09 | 0.02 | -0.19 | 0.1 | 1 | OFAMS |
| lh_isthmuscingulate_volume | -0.09 | 0.02 | -0.21 | 0.12 | 1 | OFAMS |
| lh_lateraloccipital_volume | -0.21 | -0.1 | -0.32 | 0 | 0.03 | OFAMS |
| lh_lateralorbitofrontal_volume | -0.27 | -0.16 | -0.37 | 0 | 0 | OFAMS |
| lh_lingual_volume | 0.01 | 0.13 | -0.11 | 0.88 | 1 | OFAMS |

| | | | | | | |
|---|---|---|---|---|---|---|
| lh_medialorbitofrontal_volume | -0.27 | -0.15 | -0.38 | 0 | 0 | OFAMS |
| lh_middletemporal_volume | -0.19 | -0.08 | -0.3 | 0 | 0.09 | OFAMS |
| lh_parahippocampal_volume | 0.12 | 0.26 | -0.01 | 0.08 | 1 | OFAMS |
| lh_paracentral_volume | -0.17 | -0.07 | -0.27 | 0 | 0.1 | OFAMS |
| lh_parsopercularis_volume | -0.44 | -0.34 | -0.54 | 0 | 0 | OFAMS |
| lh_parsorbitalis_volume | -0.41 | -0.3 | -0.52 | 0 | 0 | OFAMS |
| lh_parstriangularis_volume | -0.47 | -0.37 | -0.58 | 0 | 0 | OFAMS |
| lh_pericalcarine_volume | 0.2 | 0.3 | 0.09 | 0 | 0.03 | OFAMS |
| lh_postcentral_volume | -0.23 | -0.13 | -0.33 | 0 | 0 | OFAMS |
| lh_posteriorcingulate_volume | -0.13 | -0.02 | -0.25 | 0.03 | 1 | OFAMS |
| lh_precentral_volume | -0.08 | 0.03 | -0.19 | 0.16 | 1 | OFAMS |
| lh_precuneus_volume | -0.32 | -0.22 | -0.43 | 0 | 0 | OFAMS |
| lh_rostralanteriorcingulate_volume | -0.19 | -0.09 | -0.3 | 0 | 0.03 | OFAMS |
| lh_rostralmiddlefrontal_volume | -0.51 | -0.41 | -0.61 | 0 | 0 | OFAMS |
| lh_superiorfrontal_volume | -0.53 | -0.44 | -0.61 | 0 | 0 | OFAMS |
| lh_superiorparietal_volume | -0.34 | -0.23 | -0.45 | 0 | 0 | OFAMS |
| lh_superiortemporal_volume | -0.2 | -0.09 | -0.31 | 0 | 0.04 | OFAMS |
| lh_supramarginal_volume | -0.3 | -0.21 | -0.4 | 0 | 0 | OFAMS |
| lh_frontalpole_volume | -0.26 | -0.12 | -0.39 | 0 | 0.02 | OFAMS |
| lh_temporalpole_volume | 0.08 | 0.21 | -0.05 | 0.24 | 1 | OFAMS |
| lh_transversetemporal_volume | -0.29 | -0.17 | -0.41 | 0 | 0 | OFAMS |
| lh_insula_volume | -0.19 | -0.09 | -0.29 | 0 | 0.03 | OFAMS |
| rh_bankssts_volume | -0.24 | -0.12 | -0.36 | 0 | 0.01 | OFAMS |
| rh_caudalanteriorcingulate_volume | -0.06 | 0.04 | -0.16 | 0.25 | 1 | OFAMS |
| rh_caudalmiddlefrontal_volume | -0.48 | -0.38 | -0.58 | 0 | 0 | OFAMS |
| rh_cuneus_volume | -0.15 | -0.03 | -0.26 | 0.01 | 1 | OFAMS |
| rh_entorhinal_volume | 0.17 | 0.3 | 0.04 | 0.01 | 0.92 | OFAMS |
| rh_fusiform_volume | 0.16 | 0.28 | 0.04 | 0.01 | 0.83 | OFAMS |
| rh_inferiorparietal_volume | -0.18 | -0.08 | -0.28 | 0 | 0.06 | OFAMS |
| rh_inferiortemporal_volume | -0.02 | 0.09 | -0.14 | 0.67 | 1 | OFAMS |
| rh_isthmuscingulate_volume | -0.08 | 0.05 | -0.21 | 0.21 | 1 | OFAMS |
| rh_lateraloccipital_volume | -0.24 | -0.13 | -0.35 | 0 | 0 | OFAMS |
| rh_lateralorbitofrontal_volume | -0.27 | -0.16 | -0.38 | 0 | 0 | OFAMS |
| rh_lingual_volume | 0.05 | 0.18 | -0.09 | 0.5 | 1 | OFAMS |
| rh_medialorbitofrontal_volume | -0.3 | -0.19 | -0.41 | 0 | 0 | OFAMS |
| rh_middletemporal_volume | -0.15 | -0.04 | -0.26 | 0.01 | 0.93 | OFAMS |

| | | | | | | |
|---|---|---|---|---|---|---|
| rh_parahippocampal_volume | 0.12 | 0.26 | -0.01 | 0.07 | 1 | OFAMS |
| rh_paracentral_volume | -0.1 | 0 | -0.2 | 0.06 | 1 | OFAMS |
| rh_parsopercularis_volume | -0.44 | -0.34 | -0.54 | 0 | 0 | OFAMS |
| rh_parsorbitalis_volume | -0.45 | -0.34 | -0.56 | 0 | 0 | OFAMS |
| rh_parstriangularis_volume | -0.46 | -0.36 | -0.57 | 0 | 0 | OFAMS |
| rh_pericalcarine_volume | 0.1 | 0.22 | -0.03 | 0.15 | 1 | OFAMS |
| rh_postcentral_volume | -0.26 | -0.16 | -0.36 | 0 | 0 | OFAMS |
| rh_posteriorcingulate_volume | -0.11 | 0.01 | -0.23 | 0.08 | 1 | OFAMS |
| rh_precentral_volume | -0.17 | -0.06 | -0.28 | 0 | 0.32 | OFAMS |
| rh_precuneus_volume | -0.27 | -0.16 | -0.38 | 0 | 0 | OFAMS |
| rh_rostralanteriorcingulate_volume | -0.11 | -0.01 | -0.21 | 0.04 | 1 | OFAMS |
| rh_rostralmiddlefrontal_volume | -0.5 | -0.41 | -0.6 | 0 | 0 | OFAMS |
| rh_superiorfrontal_volume | -0.51 | -0.42 | -0.6 | 0 | 0 | OFAMS |
| rh_superiorparietal_volume | -0.33 | -0.22 | -0.45 | 0 | 0 | OFAMS |
| rh_superiortemporal_volume | -0.22 | -0.11 | -0.34 | 0 | 0.01 | OFAMS |
| rh_supramarginal_volume | -0.24 | -0.15 | -0.34 | 0 | 0 | OFAMS |
| rh_frontalpole_volume | -0.28 | -0.15 | -0.41 | 0 | 0 | OFAMS |
| rh_temporalpole_volume | 0.17 | 0.3 | 0.03 | 0.01 | 1 | OFAMS |
| rh_transversetemporal_volume | -0.26 | -0.13 | -0.38 | 0 | 0.01 | OFAMS |
| rh_insula_volume | -0.11 | -0.01 | -0.21 | 0.04 | 1 | OFAMS |
| Left.Amygdala | 0 | 0.12 | -0.12 | 1 | 1 | OFAMS |
| Left.Caudate | -0.4 | -0.31 | -0.49 | 0 | 0 | OFAMS |
| Left.Cerebellum.Cortex | -0.17 | -0.05 | -0.29 | 0.01 | 0.43 | OFAMS |
| Left.Hippocampus | 0.05 | 0.18 | -0.08 | 0.47 | 1 | OFAMS |
| Left.Pallidum | -0.12 | 0.01 | -0.25 | 0.08 | 1 | OFAMS |
| Left.Putamen | -0.15 | -0.04 | -0.26 | 0.01 | 0.45 | OFAMS |
| Left.Thalamus | -0.58 | -0.45 | -0.7 | 0 | 0 | OFAMS |
| Right.Amygdala | 0.01 | 0.13 | -0.11 | 0.89 | 1 | OFAMS |
| Right.Caudate | -0.3 | -0.18 | -0.42 | 0 | 0 | OFAMS |
| Right.Cerebellum.Cortex | -0.09 | 0.03 | -0.21 | 0.13 | 1 | OFAMS |
| Right.Hippocampus | 0.05 | 0.18 | -0.08 | 0.48 | 1 | OFAMS |
| Right.Pallidum | -0.24 | -0.11 | -0.37 | 0 | 0.04 | OFAMS |
| Right.Putamen | -0.13 | -0.02 | -0.24 | 0.02 | 1 | OFAMS |
| Right.Thalamus | -0.52 | -0.4 | -0.64 | 0 | 0 | OFAMS |
| lh_bankssts_volume | -0.21 | -0.16 | -0.27 | 0 | 0 | both |
| lh_caudalanteriorcingulate_volume | -0.13 | -0.09 | -0.18 | 0 | 0 | both |

| | | | | | | |
|---|---|---|---|---|---|---|
| lh_caudalmiddlefrontal_volume | -0.3 | -0.24 | -0.36 | 0 | 0 | both |
| lh_cuneus_volume | -0.08 | -0.02 | -0.14 | 0.01 | 0.78 | both |
| lh_entorhinal_volume | 0 | 0.07 | -0.07 | 0.99 | 1 | both |
| lh_fusiform_volume | -0.06 | 0.01 | -0.12 | 0.08 | 1 | both |
| lh_inferiorparietal_volume | -0.16 | -0.1 | -0.21 | 0 | 0 | both |
| lh_inferiortemporal_volume | -0.11 | -0.06 | -0.16 | 0 | 0.01 | both |
| lh_isthmuscingulate_volume | -0.15 | -0.09 | -0.21 | 0 | 0 | both |
| lh_lateraloccipital_volume | -0.15 | -0.08 | -0.21 | 0 | 0 | both |
| lh_lateralorbitofrontal_volume | -0.24 | -0.19 | -0.3 | 0 | 0 | both |
| lh_lingual_volume | -0.03 | 0.03 | -0.09 | 0.38 | 1 | both |
| lh_medialorbitofrontal_volume | -0.18 | -0.11 | -0.24 | 0 | 0 | both |
| lh_middletemporal_volume | -0.19 | -0.14 | -0.25 | 0 | 0 | both |
| lh_parahippocampal_volume | -0.01 | 0.06 | -0.08 | 0.74 | 1 | both |
| lh_paracentral_volume | -0.14 | -0.08 | -0.21 | 0 | 0 | both |
| lh_parsopercularis_volume | -0.3 | -0.25 | -0.36 | 0 | 0 | both |
| lh_parsorbitalis_volume | -0.31 | -0.25 | -0.37 | 0 | 0 | both |
| lh_parstriangularis_volume | -0.31 | -0.25 | -0.37 | 0 | 0 | both |
| lh_pericalcarine_volume | 0.08 | 0.15 | 0.02 | 0.01 | 1 | both |
| lh_postcentral_volume | -0.18 | -0.12 | -0.23 | 0 | 0 | both |
| lh_posteriorcingulate_volume | -0.21 | -0.14 | -0.27 | 0 | 0 | both |
| lh_precentral_volume | -0.14 | -0.08 | -0.2 | 0 | 0 | both |
| lh_precuneus_volume | -0.25 | -0.19 | -0.31 | 0 | 0 | both |
| lh_rostralanteriorcingulate_volume | -0.19 | -0.14 | -0.25 | 0 | 0 | both |
| lh_rostralmiddlefrontal_volume | -0.34 | -0.28 | -0.4 | 0 | 0 | both |
| lh_superiorfrontal_volume | -0.42 | -0.37 | -0.47 | 0 | 0 | both |
| lh_superiorparietal_volume | -0.25 | -0.19 | -0.31 | 0 | 0 | both |
| lh_superiortemporal_volume | -0.18 | -0.12 | -0.23 | 0 | 0 | both |
| lh_supramarginal_volume | -0.21 | -0.16 | -0.27 | 0 | 0 | both |
| lh_frontalpole_volume | -0.2 | -0.12 | -0.27 | 0 | 0 | both |
| lh_temporalpole_volume | -0.04 | 0.03 | -0.12 | 0.26 | 1 | both |
| lh_transversetemporal_volume | -0.25 | -0.19 | -0.31 | 0 | 0 | both |
| lh_insula_volume | -0.17 | -0.12 | -0.22 | 0 | 0 | both |
| rh_bankssts_volume | -0.15 | -0.09 | -0.21 | 0 | 0 | both |
| rh_caudalanteriorcingulate_volume | -0.08 | -0.02 | -0.14 | 0.01 | 0.55 | both |
| rh_caudalmiddlefrontal_volume | -0.38 | -0.33 | -0.44 | 0 | 0 | both |
| rh_cuneus_volume | -0.15 | -0.09 | -0.21 | 0 | 0 | both |

| | | | | | | |
|---|---|---|---|---|---|---|
| rh_entorhinal_volume | 0.09 | 0.16 | 0.03 | 0.01 | 0.53 | both |
| rh_fusiform_volume | 0.02 | 0.08 | -0.04 | 0.56 | 1 | both |
| rh_inferiorparietal_volume | -0.15 | -0.1 | -0.21 | 0 | 0 | both |
| rh_inferiortemporal_volume | -0.13 | -0.07 | -0.18 | 0 | 0 | both |
| rh_isthmuscingulate_volume | -0.15 | -0.09 | -0.22 | 0 | 0 | both |
| rh_lateraloccipital_volume | -0.14 | -0.08 | -0.2 | 0 | 0 | both |
| rh_lateralorbitofrontal_volume | -0.23 | -0.17 | -0.28 | 0 | 0 | both |
| rh_lingual_volume | -0.05 | 0.02 | -0.11 | 0.13 | 1 | both |
| rh_medialorbitofrontal_volume | -0.19 | -0.13 | -0.24 | 0 | 0 | both |
| rh_middletemporal_volume | -0.19 | -0.13 | -0.24 | 0 | 0 | both |
| rh_parahippocampal_volume | -0.01 | 0.06 | -0.08 | 0.84 | 1 | both |
| rh_paracentral_volume | -0.1 | -0.03 | -0.16 | 0 | 0.32 | both |
| rh_parsopercularis_volume | -0.29 | -0.24 | -0.35 | 0 | 0 | both |
| rh_parsorbitalis_volume | -0.32 | -0.26 | -0.39 | 0 | 0 | both |
| rh_parstriangularis_volume | -0.33 | -0.27 | -0.39 | 0 | 0 | both |
| rh_pericalcarine_volume | 0.04 | 0.1 | -0.03 | 0.29 | 1 | both |
| rh_postcentral_volume | -0.19 | -0.13 | -0.25 | 0 | 0 | both |
| rh_posteriorcingulate_volume | -0.18 | -0.11 | -0.24 | 0 | 0 | both |
| rh_precentral_volume | -0.17 | -0.11 | -0.23 | 0 | 0 | both |
| rh_precuneus_volume | -0.23 | -0.17 | -0.28 | 0 | 0 | both |
| rh_rostralanteriorcingulate_volume | -0.12 | -0.07 | -0.18 | 0 | 0 | both |
| rh_rostralmiddlefrontal_volume | -0.31 | -0.26 | -0.37 | 0 | 0 | both |
| rh_superiorfrontal_volume | -0.43 | -0.38 | -0.48 | 0 | 0 | both |
| rh_superiorparietal_volume | -0.21 | -0.15 | -0.27 | 0 | 0 | both |
| rh_superiortemporal_volume | -0.21 | -0.15 | -0.27 | 0 | 0 | both |
| rh_supramarginal_volume | -0.17 | -0.12 | -0.22 | 0 | 0 | both |
| rh_frontalpole_volume | -0.23 | -0.16 | -0.31 | 0 | 0 | both |
| rh_temporalpole_volume | 0.02 | 0.09 | -0.06 | 0.63 | 1 | both |
| rh_transversetemporal_volume | -0.21 | -0.14 | -0.27 | 0 | 0 | both |
| rh_insula_volume | -0.15 | -0.1 | -0.2 | 0 | 0 | both |
| Left.Amygdala | -0.12 | -0.05 | -0.19 | 0 | 0.03 | both |
| Left.Caudate | -0.29 | -0.23 | -0.34 | 0 | 0 | both |
| Left.Cerebellum.Cortex | -0.19 | -0.13 | -0.26 | 0 | 0 | both |
| Left.Hippocampus | -0.07 | 0 | -0.14 | 0.04 | 1 | both |
| Left.Pallidum | -0.12 | -0.06 | -0.19 | 0 | 0.03 | both |
| Left.Putamen | -0.16 | -0.11 | -0.22 | 0 | 0 | both |

| | | | | | | |
|---|---|---|---|---|---|---|
| Left.Thalamus | -0.31 | -0.24 | -0.38 | 0 | 0 | both |
| Right.Amygdala | -0.11 | -0.04 | -0.17 | 0 | 0.11 | both |

Values displayed as 0.00 are <0.001.

***Supplemental Table 2*. Comparison of standardized coefficients indicating the longitudinal association of regional age and brain volume between UK Biobank and MS datasets (OUH and OFAMS)**

| ROI | Z | p | ES UKB | ES MS | Data |
|---|---|---|---|---|---|
| lh caudalmiddlefrontal | 0.94 | 0.83 | -0.16 | -0.13 | OUH |
| lh lateralorbitofrontal | -1.25 | 0.11 | -0.17 | -0.21 | OUH |
| lh parsopercularis | 3.66 | 1 | -0.27 | -0.14 | OUH |
| lh parsorbitalis | -1.2 | 0.11 | -0.2 | -0.26 | OUH |
| lh parstriangularis | 3.09 | 1 | -0.28 | -0.16 | OUH |
| lh precuneus | 3.51 | 1 | -0.26 | -0.14 | OUH |
| lh rostralanteriorcingulate | -1.11 | 0.13 | -0.12 | -0.15 | OUH |
| lh rostralmiddlefrontal | 1.95 | 0.97 | -0.25 | -0.18 | OUH |
| lh superiorfrontal | -1.4 | 0.08 | -0.22 | -0.27 | OUH |
| lh superiorparietal | 2.52 | 0.99 | -0.24 | -0.14 | OUH |
| lh superiortemporal | 4.84 | 1 | -0.29 | -0.12 | OUH |
| lh supramarginal | 4.64 | 1 | -0.26 | -0.11 | OUH |
| lh frontalpole | -2.43 | 0.01 | -0.05 | -0.17 | OUH |
| lh transversetemporal | 0.47 | 0.68 | -0.18 | -0.16 | OUH |
| lh insula | -0.04 | 0.48 | -0.1 | -0.1 | OUH |
| rh caudalmiddlefrontal | -2.09 | 0.02 | -0.14 | -0.22 | OUH |
| rh lateralorbitofrontal | -1.01 | 0.16 | -0.1 | -0.13 | OUH |
| rh parsopercularis | 2.14 | 0.98 | -0.23 | -0.16 | OUH |
| rh parsorbitalis | -1.25 | 0.11 | -0.2 | -0.25 | OUH |
| rh parstriangularis | 3.26 | 1 | -0.28 | -0.16 | OUH |
| rh precuneus | 2.86 | 1 | -0.24 | -0.14 | OUH |
| rh rostralmiddlefrontal | 2.43 | 0.99 | -0.23 | -0.14 | OUH |
| rh superiorfrontal | -2.41 | 0.01 | -0.2 | -0.28 | OUH |
| rh superiortemporal | 3.14 | 1 | -0.29 | -0.17 | OUH |
| Lh Caudate | 1.71 | 0.96 | -0.23 | -0.16 | OUH |
| Lh Thalamus | 0.5 | 0.69 | -0.22 | -0.2 | OUH |
| Rh Caudate | -2.46 | 0.01 | -0.17 | -0.27 | OUH |
| Rh Thalamus | -0.27 | 0.39 | -0.27 | -0.28 | OUH |
| lh caudalmiddlefrontal | -4.62 | 0 | -0.16 | -0.43 | OFAMS |
| lh lateralorbitofrontal | -1.67 | 0.05 | -0.17 | -0.27 | OFAMS |
| lh parsopercularis | -3.05 | 0 | -0.27 | -0.44 | OFAMS |
| lh parsorbitalis | -3.56 | 0 | -0.2 | -0.41 | OFAMS |
| lh parstriangularis | -3.33 | 0 | -0.28 | -0.47 | OFAMS |
| lh precuneus | -1.11 | 0.13 | -0.26 | -0.32 | OFAMS |
| lh rostralanteriorcingulate | -1.41 | 0.08 | -0.12 | -0.19 | OFAMS |
| lh rostralmiddlefrontal | -4.98 | 0 | -0.25 | -0.51 | OFAMS |
| lh superiorfrontal | -6.51 | 0 | -0.22 | -0.53 | OFAMS |
| lh superiorparietal | -1.81 | 0.04 | -0.24 | -0.34 | OFAMS |
| lh superiortemporal | 1.63 | 0.95 | -0.29 | -0.2 | OFAMS |

| | | | | | |
|---|---|---|---|---|---|
| lh supramarginal | -0.81 | 0.21 | -0.26 | -0.3 | OFAMS |
| lh frontalpole | -2.99 | 0 | -0.05 | -0.26 | OFAMS |
| lh transversetemporal | -1.76 | 0.04 | -0.18 | -0.29 | OFAMS |
| lh insula | -1.59 | 0.06 | -0.1 | -0.19 | OFAMS |
| rh caudalmiddlefrontal | -6.38 | 0 | -0.14 | -0.48 | OFAMS |
| rh lateralorbitofrontal | -3 | 0 | -0.1 | -0.27 | OFAMS |
| rh parsopercularis | -3.83 | 0 | -0.23 | -0.44 | OFAMS |
| rh parsorbitalis | -4.4 | 0 | -0.2 | -0.45 | OFAMS |
| rh parstriangularis | -3.4 | 0 | -0.28 | -0.46 | OFAMS |
| rh precuneus | -0.48 | 0.31 | -0.24 | -0.27 | OFAMS |
| rh rostralmiddlefrontal | -5.21 | 0 | -0.23 | -0.5 | OFAMS |
| rh superiorfrontal | -6.49 | 0 | -0.2 | -0.51 | OFAMS |
| rh superiortemporal | 1.07 | 0.86 | -0.29 | -0.22 | OFAMS |
| Lh Caudate | -3.54 | 0 | -0.23 | -0.4 | OFAMS |
| Lh Thalamus | -5.52 | 0 | -0.22 | -0.58 | OFAMS |
| Rh Caudate | -2.1 | 0.02 | -0.17 | -0.3 | OFAMS |
| Rh Thalamus | -4.1 | 0 | -0.27 | -0.52 | OFAMS |
| lh caudalmiddlefrontal | -3.9 | 0 | -0.16 | -0.3 | combined |
| lh lateralorbitofrontal | -2.23 | 0.01 | -0.17 | -0.24 | combined |
| lh parsopercularis | -0.92 | 0.18 | -0.27 | -0.3 | combined |
| lh parsorbitalis | -2.86 | 0 | -0.2 | -0.31 | combined |
| lh parstriangularis | -0.66 | 0.25 | -0.28 | -0.31 | combined |
| lh precuneus | 0.39 | 0.65 | -0.26 | -0.25 | combined |
| lh rostralanteriorcingulate | -2.32 | 0.01 | -0.12 | -0.19 | combined |
| lh rostralmiddlefrontal | -2.72 | 0 | -0.25 | -0.34 | combined |
| lh superiorfrontal | -6.34 | 0 | -0.22 | -0.42 | combined |
| lh superiorparietal | -0.32 | 0.37 | -0.24 | -0.25 | combined |
| lh superiortemporal | 3.59 | 1 | -0.29 | -0.18 | combined |
| lh supramarginal | 1.52 | 0.94 | -0.26 | -0.21 | combined |
| lh frontalpole | -3.46 | 0 | -0.05 | -0.2 | combined |
| lh transversetemporal | -1.87 | 0.03 | -0.18 | -0.25 | combined |
| lh insula | -2.18 | 0.01 | -0.1 | -0.17 | combined |
| rh caudalmiddlefrontal | -7.04 | 0 | -0.14 | -0.38 | combined |
| rh lateralorbitofrontal | -3.79 | 0 | -0.1 | -0.23 | combined |
| rh parsopercularis | -2.01 | 0.02 | -0.23 | -0.29 | combined |
| rh parsorbitalis | -3.41 | 0 | -0.2 | -0.32 | combined |
| rh parstriangularis | -1.59 | 0.06 | -0.28 | -0.33 | combined |
| rh precuneus | 0.4 | 0.66 | -0.24 | -0.23 | combined |
| rh rostralmiddlefrontal | -2.4 | 0.01 | -0.23 | -0.31 | combined |
| rh superiorfrontal | -7.48 | 0 | -0.2 | -0.43 | combined |
| rh superiortemporal | 2.25 | 0.99 | -0.29 | -0.21 | combined |
| Lh Caudate | -1.86 | 0.03 | -0.23 | -0.29 | combined |
| Lh Thalamus | -2.4 | 0.01 | -0.22 | -0.31 | combined |

| | | | | | |
|---|---|---|---|---|---|
| Rh Caudate | -4.01 | 0 | -0.17 | -0.31 | combined |
| Rh Thalamus | -1.72 | 0.04 | -0.27 | -0.33 | combined |

Values displayed as 0.00 are <0.001.

***Supplemental table 3*. Overview of the OFAMS MRI scanners and acquisition protocols**

| **Protocol (N of patients)** | **1 (3)** | **2 (13)** | **3 (3)** | **4 (1)** | **5 (3)** | **6 (4)** | **7 (3)** | **8 (5)** |
|---|---|---|---|---|---|---|---|---|
| Scanner | Siemens Aera | Siemens Prisma | Siemens Skyra | Siemens Avanto | Siemens Skyra | Siemens Avanto | Siemens Aera | Philips Achieva |
| Field strength | 1.5T | 3T | 3T | 1.5T | 3T | 1.5T | 1.5T | 1.5T |
| Sequences | T1; MPRAGE T2; FLAIR | T1; MPRAGE T2; FLAIR | T1; MPRAGE T2; FLAIR | T1; MPRAGE T2; FLAIR | T1; MPRAGE T2; FLAIR | T1; MPRAGE T2; FLAIR | T1; MPRAGE T2; FLAIR | T1; FFE T2; FLAIR |
| TR (ms) | 1940 5000 | 1800 5000 | 2300 5000 | 2060 5000 | 2300 5000 | 2200 6000 | 2200 5000 | 7.6 4800 |
| TE (ms) | 2.69 335 | 2.28 386 | 2.32 387 | 3.10 340 | 2.32 387 | 2.82 358 | 2.67 335 | 3.75 338 |
| TI (ms) | 976 1800 | 900 1800 | 900 1800 | 1100 1800 | 900 1800 | 900 2200 | 900 1800 | 1660 1650 |
| Flip angle (¬∞) | 8 120 | 8 120 | 8 120 | 15 120 | 8 120 | 8 120 | 8 120 | 8 90 |
| Voxel size (mm) | 1.00x0.98x0.98 1.00x1.00x1.00 | 1.00x1.00x1.00 | 1.00x1.00x1.00 | 1.00x1.00x1.00 | 0.9x0.94x0.94 0.9x0.45x0.45 | 1.00x0.49x0.49 1.00x0.51x0.51 | 0.90x0.94x0.94 0.90x0.45x0.45 | 1.00x0.98x0.98 1.00x1.00x1.00 |

| **Protocol (N of patients)** | **9 (12)** | **10 (2)** | **11 (6)** | **12 (1)** | **13 (2)** | **14 (8)** | **15 (3)** |
|---|---|---|---|---|---|---|---|
| Scanner | Philips Achieva | Siemens Prisma | Philips Ingenia | Toshiba MRT200 SP3 | Philips Ingenia | Siemens Prisma | Philips Achieva |
| Field strength | 3T | 3T | 1.5T | 1.5T | 3T | 3T | 1.5T |
| Sequences | T1; FFE T2; FLAIR | T1; MPRAGE T2; FLAIR | T1; FFE T2; FLAIR | T1; FFE T2; FLAIR | T1; FFE T2; FLAIR | T1; MPRAGE T2; FLAIR | T1; FFE T2; FLAIR |
| TR (ms) | 8.04 5000 | 1800 5000 | 25 4800 | 13.5 1160 | 11.11 4800 | 1800 5000 | 7.1 4800 |
| TE (ms) | 3.68 386 | 2.28 385 | 9.21 367 | 5.50 105 | 6.29 296 | 2.26 387 | 2.2 307 |
| TI (ms) | 900 1800 | 900 1800 | 1660 2300 | 1650 906 | 1800 | 1800 | 1660 |
| Flip angle (¬∞) | 8 90 | 8 120 | 30 90 | 20 90 | 8 90 | 8 120 | 8 90 |
| Voxel size (mm) | 1.00x0.98x0.98 1.00x0.98x0.98 | 1.00x0.46x0.46 0.50x0.73x0.73 | 1.00x0.50x0.50 1.00x0.50x0.50 | 1.00x0.94x0.94 0.50x0.74x0.74 | 0.39x0.65x0.39 0.47x4.0x0.47 | 0.90x0.67x0.67 0.56x0.98x0.98 | 1.00x1.000x1.00 1.00x1.000x1.00 1.00x1.00x1.00 0.76x1.00x1.00 |

### Supplemental Figures

***Supplemental Figure 1*. Volume changes in relapsing-remitting multiple sclerosis and age-associations in healthy controls using scanner as fixed effect instead of Combat harmonisation.**

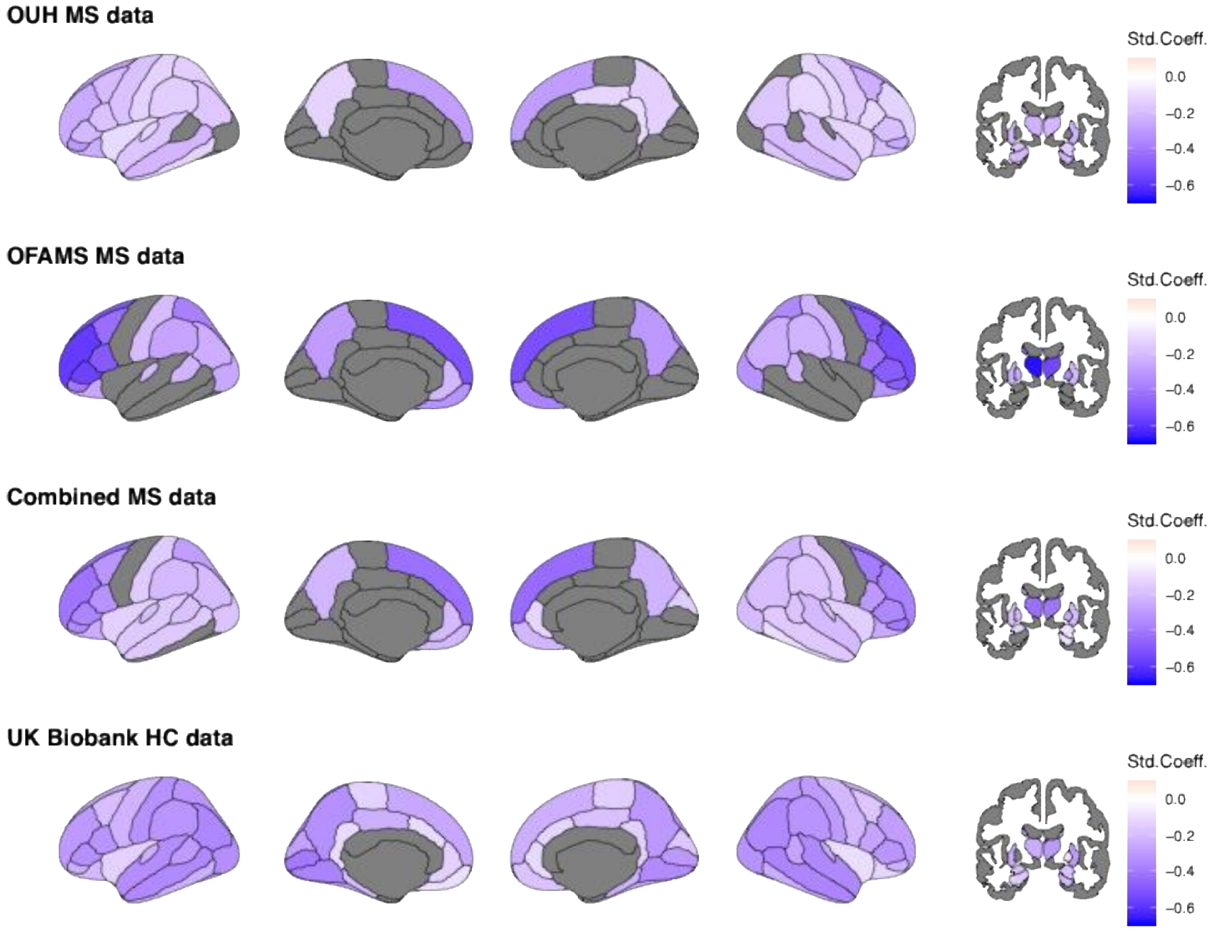


Top row: ageing effects in the OUH sample (baseline N = 165). Second row: ageing effects in the OFAMS sample (baseline N = 85). Third row: ageing effects in both MS samples combined. Fourth row: ageing effects in the UK Biobank sample of healthy controls (N = 1,586). Grey regions indicate Bonferroni-corrected $p \geq 0.05$.

Note that, besides stronger significance, indicated by lower p-values, particularly the thalamus was highlighted as a significantly degenerating region in pwMS. The thalami degenerated also faster than in UK Biobank (Supplemental Figure 2) and were significantly related to EDSS (Supplemental Note 2). Overall, more regions presented significant degeneration which was replicable across MS dataset.

***Supplemental Figure 2*. Volume changes in MS compared to 20 years older healthy controls using scanner as fixed effect instead of Combat harmonisation.**

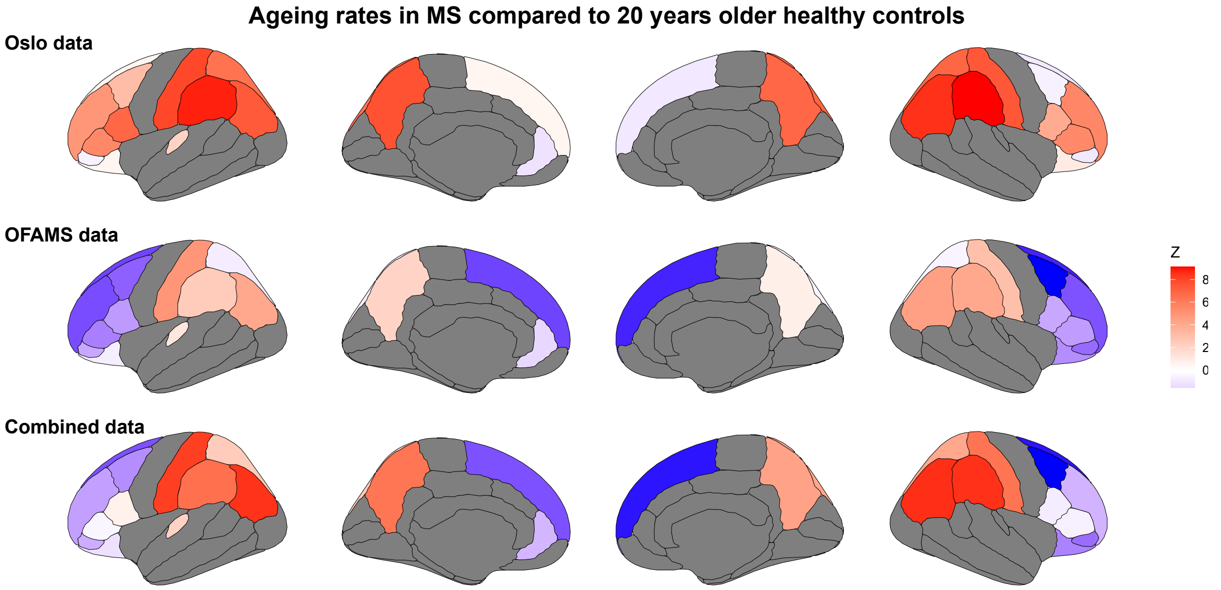


The regions considered for this analysis were selected based on cross-sample replicated brain changes, with brain regions excluded from these analyses being marked in grey. Blue colours indicate faster ageing in MS, white colour indicates no difference, and red colour indicates faster ageing in older healthy controls. The top two rows indicate comparisons between standardised regression coefficients of single MS datasets and UK Biobank data. The bottom row indicates such comparisons between coefficients from combined MS data and UK Biobank healthy controls. Note, the presented significance level is not adjusted for multiple comparisons. Only cortical areas are displayed due to a small number of significant subcortical areas, including the thalamus. The findings correspond closely to the findings reported in the main text for the OFAMS and combined sample. For the OUH sample, the effects are smaller.

### Supplemental Notes

#### *Supplemental Note 1:* Associations between clinical scores and regional atrophy when analysing both samples together (for greater statistical power)

Analysing both samples together yielded multiple significant associations of EDSS with subcortical volumes (Figure below, right panel), of which the thalami (left: $\beta_{EDSS}$=-0.09 [-0.05, -0.13], $p_{Bonferroni}$<0.001; right: $\beta_{EDSS}$=-0.10 [-0.05, -0.14], $p_{Bonferroni}$=0.005) were among the regions with significant matter volume loss. Additional significant regions included the hippocampi (left: $\beta_{EDSS}$=-0.16 [-0.10, -0.23], $p_{Bonferroni}$<0.001; right: $\beta_{EDSS}$=-0.12 [-0.07, -0.18], $p_{Bonferroni}$=0.001), left putamen ($\beta_{EDSS}$=-0.09 [-0.05, -0.14], $p_{Bonferroni}$=0.005), and right lateral occipital lobe ($\beta_{EDSS}$=-0.08 [-0.04, -0.12], $p_{Bonferroni}$=0.026).

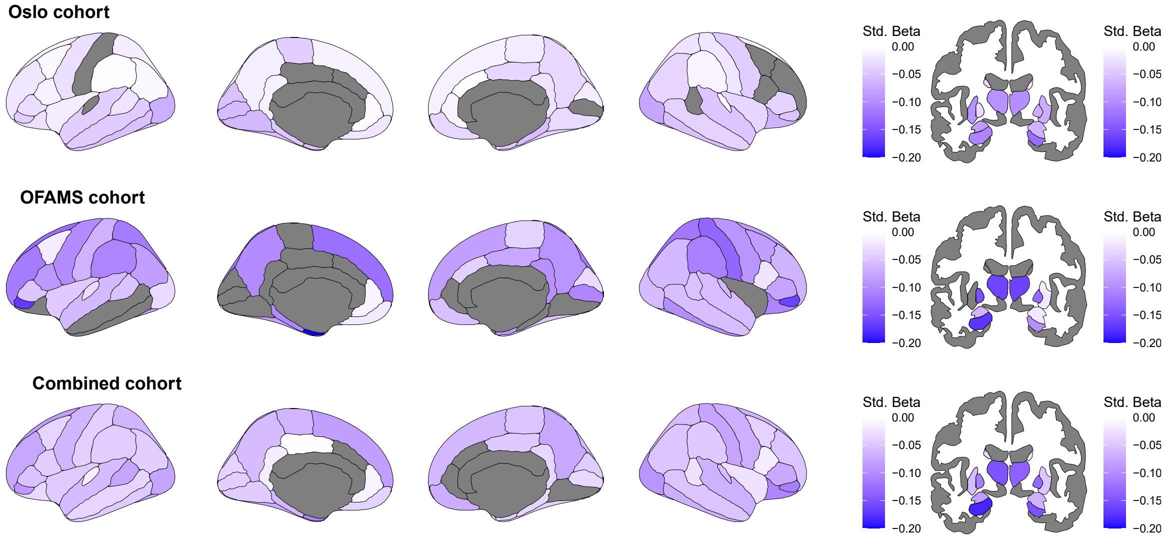


EDSS - brain volume associations across time in both MS samples.

Despite analysing both cohorts together, no effects of brain volume on fatigue or PASAT survived multiple comparison. Assessing only uncorrected p-values within significantly degenerating areas highlighted right caudal middle frontal ($\beta_{fatigue}$=-0.09 [-0.004, -0.18], p=0.041) and right lateral orbitofrontal volume ($\beta_{fatigue}$=-0.13 [-0.03, -0.23], p=0.008) associations with fatigue, and right lateral orbitofrontal volume ($\beta_{PASAT}$ =0.11 [0.005, 0.23], p=0.040), left caudate ($\beta_{PASAT}$=0.09 [0.003, 0.18], p=0.003), and right thalamus ($\beta_{PASAT}$ =0.13 [0.02, 0.25], p=0.023) correlations with PASAT.

***Supplemental Note 2.*** **Reanalysis of clinical score associations with brain volumes using scanner as a fixed effect instead of Combat harmonisation.**

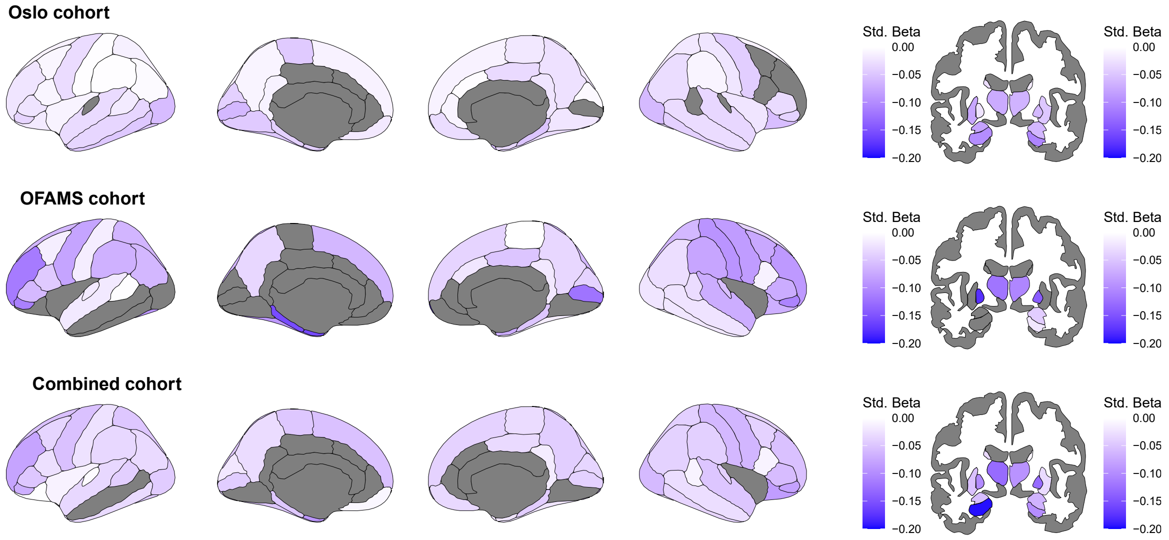


EDSS associations resemble the findings in the main text with slightly smaller effects in OUH and slightly stronger effects in the OFAMS data. Note however that the effects in the OFAMS data do not pass the significance level of 0.005 defined in-text. Hence, the replicable patterns presented in the main text could not be shown when addressing scanner effects as simple fixed effects. The same holds true for PASAT and fatigue associations with regional brain volumes. Note, that OFAMS is a multicentre study requiring harmonisation for non-biological effects.

***Supplemental Note 3*. Post-hoc power analysis.**

Post-hoc power simulations were obtained for the reported effects, here, focussing on the effects highlighted in the manuscript. The power of ageing effects of pars orbitalis, thalami and superior frontal cortices approximated 100%. Associations of EDSS with the left thalamus was 99% [94.55, 99.97] in OUH and 70% [60.02, 78.76] in OFAMS, and in the right thalamus 99% [94.55, 99.97] and 69% [58.97, 77.87], respectively. The power for associations between the left hippocampus and EDSS were 98.00% [92.96, 99.76] in PUH and 73.00% [63.20, 81.39] in OFAMS. Power was similarly low across samples for the right pallidum, for OUH: 57.00% [46.71, 66.86] and OFAMS: 63.00% [52.76, 72.44].

Finally, the power for associations between EDSS changes and changes in left entorhinal volumes were 60.00% [49.72, 69.67] in OUH and 81.00% [71.93, 88.16] in OFAMS. Whole brain grey matter and EDSS change associations were well-powered in both samples both = 100.0% [96.38, 100.0].

***Supplemental Note 4*. Data Acquisition of the OUH MS sample**

All pwMS were scanned at several time points between January 2012 and August 2017 in a study setting, using the same 1.5 T scanner (Avanto, Siemens Medical Solutions; Erlangen, Germany) equipped with a 12-channel head coil. Structural MRI data were collected using a 3D T1-weighted MPRAGE (Magnetization Prepared Rapid Gradient Echo) sequence, with the following parameters: TR (repetition time)/TE (echo time)/flip angle/voxel size/FOV (field of view)/slices/scan time/matrix/time to inversion = 2,400 ms/3.61 ms/8°/1.20 × 1.25 × 1.25 mm/240/160 sagittal slices/7:42 min/192 × 192/1,000 ms. The MRI sequence was kept identical during the scanning period. FLAIR (Fluid attenuation inversion recovery), T2 and pre- and post-gadolinium 3D T1 sequences were attained and used for neuroradiological evaluation.

Fifty-eight of the people with MS were also scanned at OUH at 3T with a GE 750 Discovery MRI scanner using a 32-channel head coil at time point 3 between August 2016 and June 2017. During the same week they were scanned at the 1.5 T scanner for time point 3. HCs were scanned solely on the 3 T scanner at one time point to provide cross-sectional data. Structural MRI data were collected using a 3D high-resolution IR-prepared FSPGR (fast spoiled gradient echo) T1-weighted sequence (3D BRAVO) with the following parameters: TR (repetition time)/TE (echo time)/flip angle/voxel size/FOV (field of view)/slices/scan time = 8.16 ms/3.18 ms/12°/1 × 1 × 1 mm/256 × 256 mm/188 sagittal slices/4:42 min.

***Supplemental Note 5*. Comparison of key ageing regions between MS and HC**

As a comparison to the ageing effects in the MS samples, we also examined the effect of age on regional brain volumes in HCs using the longitudinal sample (UK Biobank), and the cross-sectional lifespan sample, presenting comparably smaller effects on the *superior frontal cortex* (UK Biobank left: $\beta_{age}$=-0.22 [-0.19, -0.25], $p_{Bonferroni}$<0.001, right: $\beta_{age}$=-0.20 [-0.17, -0.23], $p_{Bonferroni}$<0.001; Lifespan sample left: : $\beta_{age}$=-0.27 [-0.26, -0.28], $p_{Bonferroni}$<0.001, right: $\beta_{age}$=-0.25 [-0.24, -0.25], $p_{Bonferroni}$<0.001), *pars orbitalis* (UK Biobank left: $\beta_{age}$=-0.20 [-0.17, -0.24], $p_{Bonferroni}$<0.001, right: $\beta_{age}$=-0.20 [-0.16, -0.23], $p_{Bonferroni}$<0.001; Lifespan sample left: : $\beta_{age}$=-0.30 [-0.29, -0.31], $p_{Bonferroni}$<0.001, right: $\beta_{age}$=-0.30 [-0.29, -0.31], $p_{Bonferroni}$<0.001), and *thalami* (UK Biobank left: $\beta_{age}$=-0.22 [-0.19, -0.25], $p_{Bonferroni}$<0.001, right: $\beta_{age}$=-0.27 [-0.24, -0.30], $p_{Bonferroni}$<0.001; Lifespan sample left: $\beta_{age}$=-0.24 [-0.23, -0.25], $p_{Bonferroni}$<0.001, right: $\beta_{age}$=-0.33 [-0.32, -0.34], $p_{Bonferroni}$<0.001).

### References


1. Lovestone, S. *et al.* AddNeuroMed--the European collaboration for the discovery of novel biomarkers for Alzheimer's disease. *Ann. N. Y. Acad. Sci.* **1180**, 36–46 (2009).
2. Van Essen, D. C. *et al.* The Human Connectome Project: A data acquisition perspective. *NeuroImage* **62**, 2222–2231 (2012).
3. Nooner, K. B. *et al.* The NKI-Rockland sample: a model for accelerating the pace of discovery science in psychiatry. *Front. Neurosci.* **6**, 152 (2012).
4. Alfaro-Almagro, F. *et al.* Image processing and Quality Control for the first 10,000 brain imaging datasets from UK Biobank. *NeuroImage* **166**, 400–424 (2018).
